# Evaluating the policy of closing bars and restaurants in Cataluña and its effects on mobility and COVID19 incidence

**DOI:** 10.1101/2021.12.03.21267172

**Authors:** Matthew Smith, Miguel Ponce-de-Leon, Alfonso Valencia

## Abstract

The world has gone through unprecedented changes since the global pandemic hit. During the early phase of the pandemic, the absence of known drugs or pharmaceutical treatments forced governments to introduce different policies in order to help reduce contagion rates and manage the economic consequences of the pandemic. This paper analyses the causal impact on mobility and COVID19 incidence from policy makers in Cataluña, Spain. We use annonimized phone-based mobility data together with reported incidence and apply a series of causal impact models frequently used in econometrics and policy evaluation in order to measure the policies impact.. We analyse the case of Cataluña and the public policy decision of closing all bars and restaurants down for a 5 week period between the 2020-16-10 to 2020-23-11. We find that this decision led to a significant reduction in mobility. It not only led to reductions in mobility but from a behavioural economics standpoint we highlight how people responded to the policy decision. Moreover, the policy of closing bars and restaurants slowed the incidence rate of COVID19 after a time lag has been taken into account. These finding are significant since governments worldwide want to restrict movements of people in order to slow down COVID19 incidence without infringing on their rights directly.

## Introduction

The world has gone through unprecedented changes since the global pandemic hit. Peoples lives have changed drastically with stay-at-home orders, curfews and strict lock-downs. The highly contagious nature of the COVID19 virus, together with little to no regulations on restrictions on global mobility during the early periods of the pandemic resulted in the spread of the virus to every corner of the world. During the early phase of the pandemic, the absence of known drugs or pharmaceutical treatments forced governments to introduce different policies in order to help reduce contagion rates and manage the economic consequences of the pandemic. These policies, also referred to as non-pharmaceutical interventions (NPIs) include, among other things, closing of national borders, travel restrictions, restrictions on public gatherings, restricting the capacity in bars, restaurants, theaters and emergency investment into healthcare systems and new forms of social welfare provisions. Different stages of lockdowns have been introduced by governments across the world, (1) studied the impact of 130 NPIs across 130 countries, (2) construct a policy indicator *Oxford COVID19 Government Response Tracker* (OxCGRT) which provides a systematic way to track government responses to COVID19 over time by taking 20 indicators such as school closures and travel restrictions for more than 180 countries. Since public health experts are learning in real time, there is debate in politics over the level of response which should be pursued and the timescale to implement and roll-back different responses. Many NPIs have been proven to have a positive impact on controlling the spread of COVID19, (3) analysed how the implementation of NPIs, along with climatic, social and demographic factors all affected the initial growth of COVID19. They state that government introduced NPIs do not explain the growth in COVID19 cases and that the growth rate in cases is explained by demographic, climatic and social variables. (4) studied the role of public attention to COVID19 using Google search data, finding that countries with higher levels of public attention are more likely to implement NPIs and the extent to which a government is willing to introduce NPIs is dependent on the countries institutional quality. Moreover, it is important to evaluate the impact of NPIs in order to gain a deeper insight into both the positive effects, in terms of pandemic control and the negative effects, in terms of the economy.

Population density may also form an important characteristic in COVID19 infection rates, maintaining safe distances is more difficult in areas with higher population densities (5). Other studies have found that there has been moderate associations between population densities and the spread of COVID19 (6), (7) and (8). Moreover, other literature did not find the same results suggesting that more densely populated areas have better access to health care and increased social distancing policies (9). More densely populated areas may tend to experience an outbreak earlier than more sparsely populated areas, however, (10) found no evidence that population density is linked with COVID19 cases.

In order to help reduce the spread of COVID19, policy makers throughout the world have sought to place restrictions on local populations. These reductions on human interactions should lead to reductions in person to person contagion but it also comes at a high political, social and economic cost. (11) studied the impact of COVID19 on the U.K. economy by linking a macroeconomic model to an epidemiological model. They state that applying mitigation strategies for 12 weeks reduced fatalities by 29% but the cost tot he economy was 13.5% of GDP of which 2.9% is attributable to labour lost from parents staying at home due to school closures and 8.8% is attributable to business closures. Moreover, the shut down of businesses in Europe was estimated to reduce GDP by 3% per month (12). Therefore it is important to find the right balance between public health, societal and economic costs.

This paper address the problem of evaluating non-pharmaceutical interventions (NPI) and public policy decisions during the COVID19 pandemic in Spain. Specifically, we have focused on evaluating a policy brought in to help slow the spread of COVID19 incidence during a time when Cataluña was experiencing a peak in the number of daily new COVID19 cases. The policy forced the closure of bars and restaurants, only allowing takeaway services along with a maximum capacity in shopping centres reduced to 30%, fitness clubs, cinemas and theatres capacity restricted to 50%, the suspension of all non-professional sporting activities and face-to-face teaching was also suspended. These measures were originally supposed to last two weeks but remained in place for five weeks in total. At the time, local health authorities were seeing a total of 12,211 new cases between October 4th 2020 and October 10th 2020 and rising. Hospitals in the region had more than 1,000 COVID19 hospitalisations, 189 of whom were in intensive care units (ICUs), the R rate (measuring how many people a positive case may infect) was at 1.3. This prompted the Catalan government to implement measures to reduce social contact which is the main driver in COVID19 outbreaks.

The introduction of the policy was brought on by the regional government of Cataluña and not that of the central government of Spain. The ministry of health for Spain (Ministerio de Sanidad, Consumo y Bienestar Social) agreed in September of 2020 that regions in Spain (with more than 100,000 inhabitants) must be confined should the following thresholds be exceeded, a 14-day cumulative number of COVID19 cases above 500, ICU occupancy greater that 35% and a positivity rate greater than 10%. At the time Cataluña had 7% of hospital beds occupied by COVID19 patients with an ICU occupancy of 19.5%. Madrid on the otherhand had 21% of hospital beds occupied by COVID19 patients and 38% ICU occupancy. It is important to note that in Spain, regional governments take the primary decisions on public heath initiatives with the central government stepping in when necessary. For example, at the time, the regional government of Madrid lack of action prompted the central government of Spain to declare a state of alarm in the region, overriding the regional government and putting in place a perimetral confinement for Madrid’s municipalities in order to bring infection rates down. The policy under evaluation in this paper was introduced by the Catalan government and not the central government as was the case with Madrid.

Using annonimized phone-based mobility data we analyse the change in mobility patterns after the introduction of a policy made in Cataluña of closing down bars and restaurants in order to try and slow the spread of COVID19 infection rates. For this propose, we collected anonymous mobile phone data based on trips and construct origin destination (OD) matrices for each day. Each OD matrix accounts for the number of trips from a given mobility zone into another mobility zone, where individual mobility zones make up a combination of municipalities and districts across Spain. Using the OD matrices we compute four types of mobility indexes *incoming, outgoing, internal* and *total*. The *incoming* and *outgoing* mobility types are trips coming to and from a given mobility zone, *internal* concerns the movement of trips within a given mobility zone and *total* refers to a summation of the three mobility types, a more detailed explanation is left to the methodology section. To assess the effect of the analyzed policy on the mobility, we apply a series of causal inference models which are useful in quantifying the impact of public policy decisions or NPI’s. Most notably, we apply an Ordinary Least Squares (OLS) model with policy controls, a difference-in-difference model and a Bayesian structural time-series model. We find that all models agree that the introduction of the policy helped to reduce mobility levels. We find that this policy affected the behaviour of people and lead to significant reductions in mobility. That is, people responded to the NPI policy by travelling less across mobility zones but did not alter their behaviour within their own mobility zone.

## 1 Literature Review

This section discusses relevant literature. We first break the section down to related literature using interrupted time series for policy decisions. Secondly, we link mobility with public policy decision making and finally we discuss how mobility has been linked to COVID19 incidence.

### 1.1 Interrupted time series

Interrupted time series models are being increasingly applied when analysing the impact of public health interventions. (13) applied Box-Tiao intervention analysis to analyse the effects of the introduction of U.S. legislation requiring the use of mandatory seat belts across 8 states between 1976 and 1986. (14) applied interrupted time series modeling to study the association between the introduction of helmet legislation and admissions to hospital for cycling related head injuries between 1994 and 2003 in Canada. (15) used interrupted time series to asses the effect of U.K. legislation of reducing package sizes of paracetamol on deaths from paracetamol poisoning between 1998 and 2009. (16) used interrupted time series to study the effect of the introduction of 20 mph (32 km an hour) traffic speed zones on road collisions, injuries and fatalities in London between 1986 and 2006. (17) used interrupted time series to analyse the association between the 2008 financial crisis and suicide rates in Spain, using data between 2005 and 2010. Interrupted time series models have long be used to analyse public policy and time series related events in economics and provides a suitable methodology for analysing policy events related to COVID19.

### 1.2 Mobility and public policy

(18) used Google Community Mobility Data to analyses mobility changes from a baseline mobility level (pre-pandemic). They found three distinct patterns of societal reaction to social restrictions. In Australia (which implemented a near complete lockdown) people did not go to their workplaces and stayed at home. In Sweden (which implemented relaxed lockdown with preserved workplace activity) the change in workplace mobility was smaller suggesting preservation of workplace activity. South Korea (which implemented minimal lockdown and preservation of workplace activity) changes in workplace mobility was even smaller. These finding suggest that populations respond directly to governmental interventions and highlights the important role of government decisions on day to day lives of its populations. (19) analysed public policy effects on driving, transit and walking mobility behaviour, finding that they dropped to lower levels in Canada than the U.S. during March 2020 and show strong evidence that policy effects mobility behaviour. (20) used a difference-in-difference approach and mobility data to study the causal impact of policies finding that statewide stay-at-home orders had the strongest causal impact on reducing social interactions and affects both mobility patterns and subsequent infection rates. (21) quantifies the effect of U.S. state reopening policies on daily mobility levels, finding that four days after reopening mobility increased between 6% to 8%. Public policy decision therefore can have a direct effect on the behavioural patterns of inhabitants mobility in regions directly affected by the policy.

### 1.3 Mobility and its relation to COVID19 incidence

(22) show that mobility patterns are strongly correlated with decreased COVID-19 case growth rates in the USA. They collected data from 1st January 2020 to 20th April 2020. They define a Mobility Ratio (MR) to quantify the change in mobility patterns when compared to a baseline day before the pandemic when travel patterns were stable. They use this as a proxy for social distancing such that when an individual makes fewer trips, they interact less. They link mobility data with data on cases and construct a COVID-19 Growth Ratio (GR) in order to capture the complex and time-dependent dynamics between mobility and cases. They show a statistically significant correlation between their socially distancing metric and reductions in COVID-19 growth rates, showing that the effect of social distancing on case growth is not likely to be noticed for at least 9-12 days after implementation. Moreover, (23) extended their own research to include more regions and expanded the time horizon from 16th March 2020 to 16th September 2020. They found that the linear association between mobility and case growth rates previously observed is absent after April 2020 and that mobility has a less significant role in the transmission of COVID-19 than other adopted behavioural changes and NPIs such as wearing face masks, hand-washing, maintaining physical distance, avoiding large gatherings and closing schools. The strong association revealed in March-April is related to the adoption of NPIs in parallel and after an introduction of varying policies and changes in individuals mobility behaviours confound the role of mobility. They conclude that using mobility data alone is likely to result in inaccurate models and forecasts and that there are more critical factors than mobility for controlling COVID-19. (24) also identified a strong correlation between decreased mobility and reduced COVID-19 case growth during the period 27th March 2020 and 20th April 2020. They found that when they extended their time-horizon between 21st April 2020 and 24th May 2020 and later 25th May 2020 to 22nd July 2020, that there was only a weak correlation between daily distance difference and case growth. They find that mobile phone data only captures a small component of the behaviours associated with social distancing and reduced case growth rates and that other NPIs such as wearing masks, maintaining distance are likely to be more important than mobility alone.

(25) used mobile phone data and modelled the relationship between mobility inflow and infections across counties in the U.S. between March 1 to June 9 2020. They found that travel between counties decreased 35% after the U.S. entered a lockdown but recovered rapidly once the lockdown started to ease. Using a system of equations they find a strong and positive relationship between mobility and the number of infections across counties with an average coefficient of 0.243, that is, a 10% increase in mobility is associated with a 2.34% increase in infections a week later. (26) analysed mobility data in China to track population outflows from Wuhan between January 1st and January 24th 2020 and linked it with COVID-19 infection counts by location. They find a strong correlation between total population flow and the number of infections across regions. (27) construct network maps of hourly movements of people to and from non-residential locations and apply a meta-population SEIR model with susceptible (S), exposed (E), infectious (F) and recovered (R) compartments which tracks the trajectories of infections. (28) used aggregated mobile phone data to build a SEIR model for the city Shenzhen, China. They simulate how the spread of COVID-19 changes when the type and magnitude of mobility restrictions varied. They found that reducing mobility by 20% delayed the epidemic peak by around 2 weeks with the peak incidence decreasing to 33%, a 40% reduction in mobility was associated with a delay of 4 weeks and reduced the peak by 66% and a 60% reduction was associated with a delayed peak of 14 weeks, decreasing the magnitude by 91%.

## 2 Results

This section discusses the results in which we asses the effect of NPIs on mobility. We compare the mobility in Cataluña with that of other regions in Spain where other policies or no policies have been applied. We apply an OLS regression, difference-in-difference and Bayesian structural time series model in order to try and capture the causal effect of this policy intervention. We first report some analysis of the mobility data and OD matrices for Cataluña and Madrid, two of the most populous regions in Spain and where one region introduced the policy whereas the other did not. We then report the main results of the paper from the different econometric models. Finally, we try to link the policy to reductions in COVID19 growth rates.

### 2.1 Analysis of mobility

Figure 2 shows a time series plot of the total number of trips for Cataluña and Madrid. The policy came into effect right in the center of the time series and lasted until the end of the time series (shaded as yellow) thus we see a shift down in the number of trips for Cataluña after the introduction of the policy and it remained down for the duration of the policy. Contrasted with Madrid in which we see no shift downward, which is, as we would expect since, Madrid did not introduce the same policy as Cataluña.

**Figure 1.**
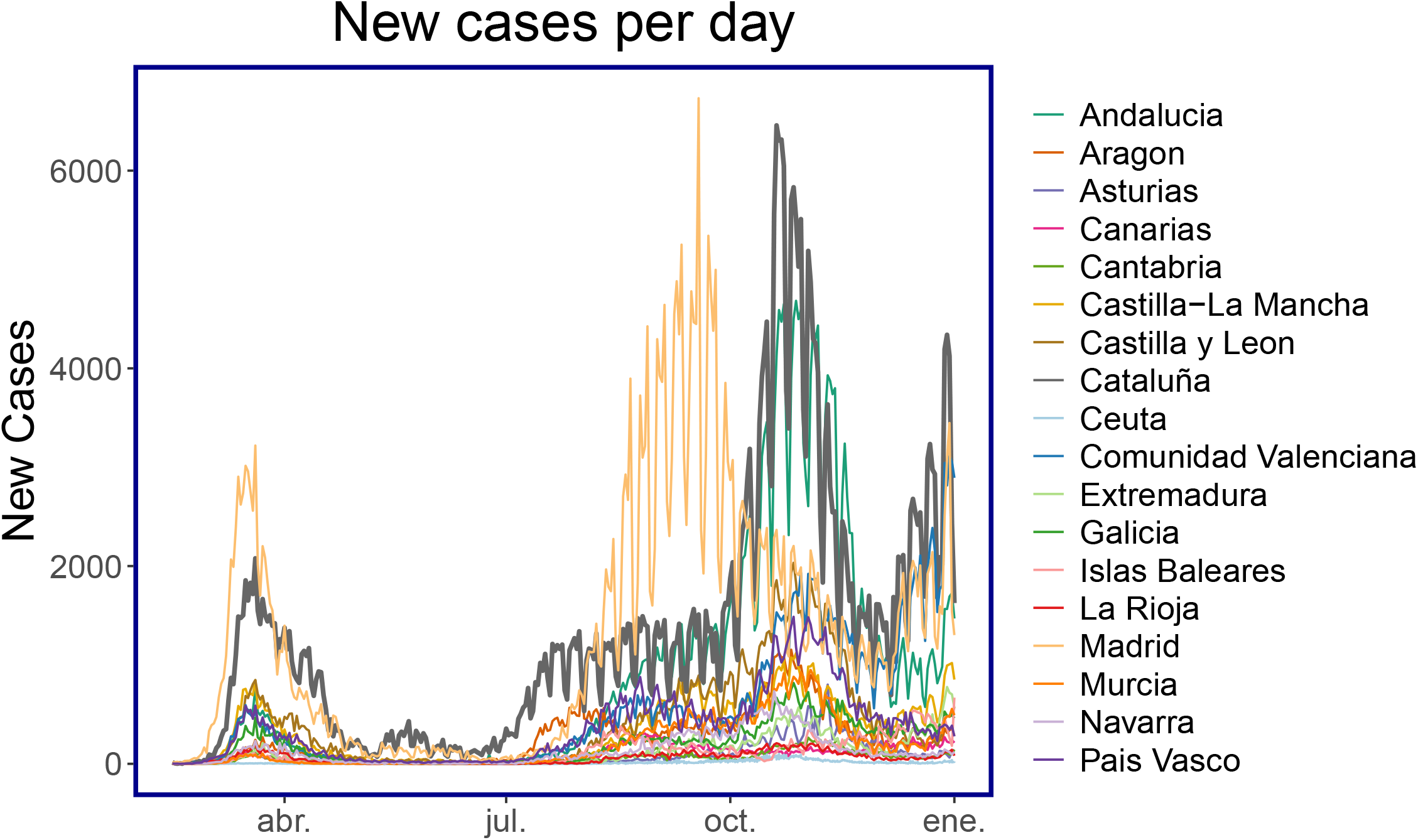
New cases per day for each CCAA: Number of COVID19 cases for each autonomous community (CCAA) in Spain. Madrid experienced its second peak before the rest of Spain, with Cataluña (as the bold line) experiencing its second peak just after. The policy was a direct response to control the outbreak of the second peak in Cataluña.

**Figure 2.**
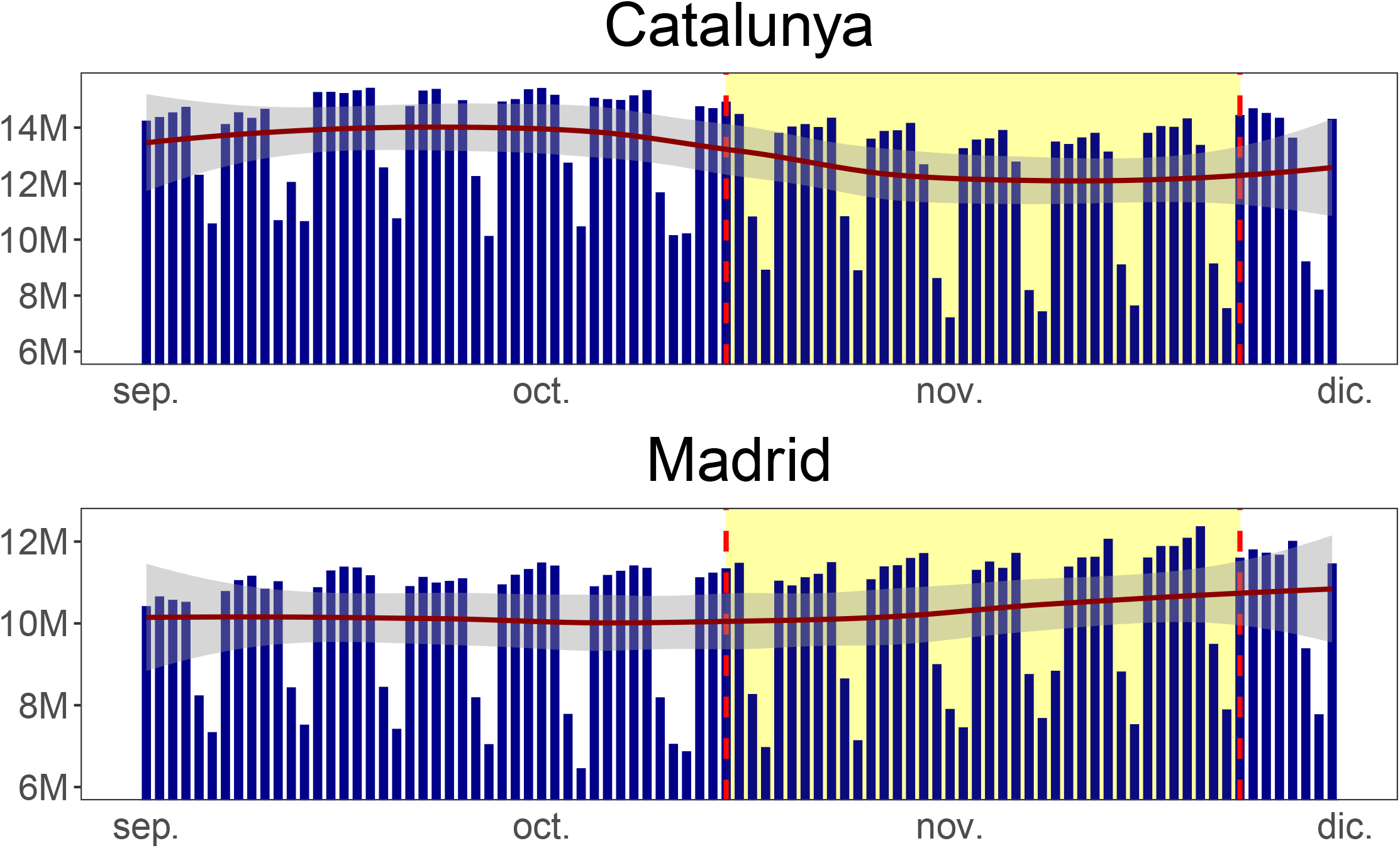
Number of trips in Cataluña and Madrid: The shaded region corresponds to the time duration when the bars and restaurants were closed in Cataluña. The pre-policy trend for both regions are relatively flat and parallel whereas the post-policy trend drops for Cataluña and not Madrid since the policy was only introduced in Cataluña.

Figure 3 shows the mobility matrices for Cataluña and Madrid for each day for incoming trips into MITMA regions. The darker colours represent higher mobility and the lighter regions lower mobility. The plot has been arranged to show MITMA regions with overall higher mobility on the right and MITMA regions with overall lower mobility on the left. There are distinct periods of higher and lower mobility, the higher mobility corresponds to weekdays and thus this mobility incorporates people travelling to and from work and the lighter periods correspond to weekends where people travel less. Additionally, there are other days of lower mobility. At point (A) in Figure 3 there are 3 days of lower mobility, 2 of these days are a weekend and the 3rd day is a public holiday in Cataluña falling on a Friday, 2020-09-11 (Fiesta Nacional de Cataluña). The same region in the Madrid plot does not show this reduced mobility since the public holiday is only specific to Cataluña and thus not Madrid, therefore people in Madrid went to work on this day. There are other public holidays in this data, specific to Madrid which fell on Monday, 2020-11-01 (Fiesta de Todos los Santos) and Monday, 2020-11-09 (Fiesta de la Almudena) which are shown in the mobility matrix data as (C) and (D) respectively. Finally, there is a public holiday which is celebrated across the whole of Spain, on the 2020-10-12 (Fiesta Nacional de España) denoted as point (B). Therefore, mobility for this day was reduced in both Cataluña and Madrid along with other Autonomous Communities. The next sections reports the main findings of this study.

**Figure 3.**
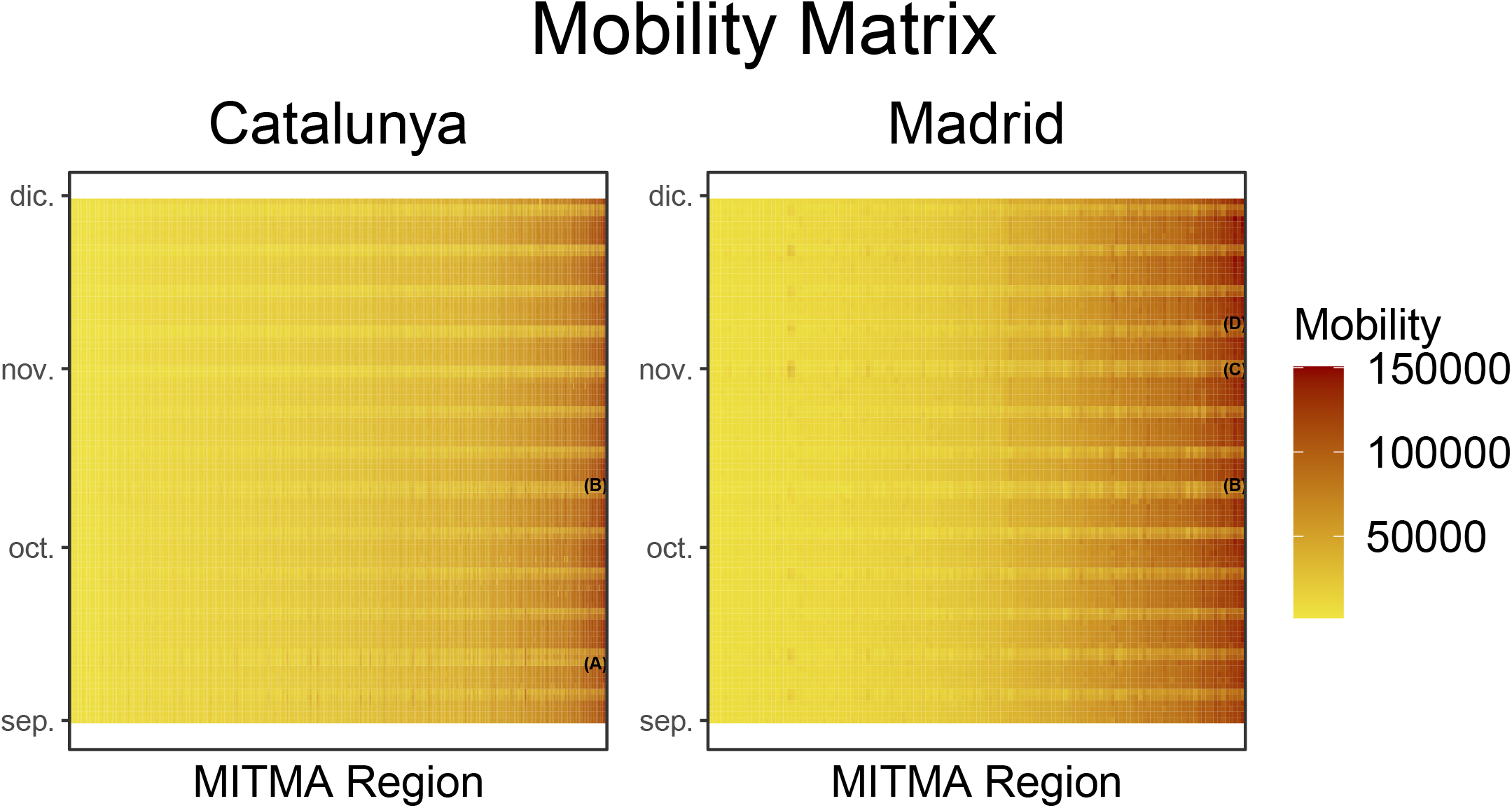
Number of incoming trips in Cataluña and Madrid: Consider annotation (A) which is displayed in the Cataluña figure but not in the Madrid figure. There was a reduction in mobility at this point (indicated by a lighter shade of yellow) because there was a public holiday in Cataluña, there was no public holiday in Madrid on this date. Moreover, annotation (B) shows a reduction in mobility for both Cataluña and Madrid. On this date there was a national holiday annd thus both regions mobility were reduced.

### 2.2 Linear regression

We first applied a series of linear regression models. Firstly, using all MITMA zones in Cataluña and then secondly, we aggregate the data in order to analyse the effect for the whole of Cataluña.

#### 2.2.1 Linear regression across all MITMA zones (Cataluña)

Table 1 reports the OLS regression results for all MITMA regions in Cataluña. There is a statistically significant drop of 15.4% in mobility on weekends whereas there is just a 3.9% drop in mobility during the weekdays. The significant drop on the weekends may suggest that people are socialising less and are remaining at home and that they do not substitute their socialising at bars and restaurants with other socialising activities such as going to the cinema, park etc. thus, We see a reduction in activities which imply the movement of people. We expect to see less of a drop during the weekdays than on weekends since people still need to go to work. However, the 3.9% drop during the weekdays may suggest that people are socialising less in bars and restaurants after work and are going straight home. After the policy impact there appears to be a sustained drop in daily mobility of 6.9% after controlling for weekend effects and therefore the policy caused a shock to mobility and then leveled out, indicated by the trend of 0.1% afterwards. Therefore, using a regression model, we have aimed to quantify the drop in mobility as seen in the Cataluña panel of Figure 2.

**Table 1.**
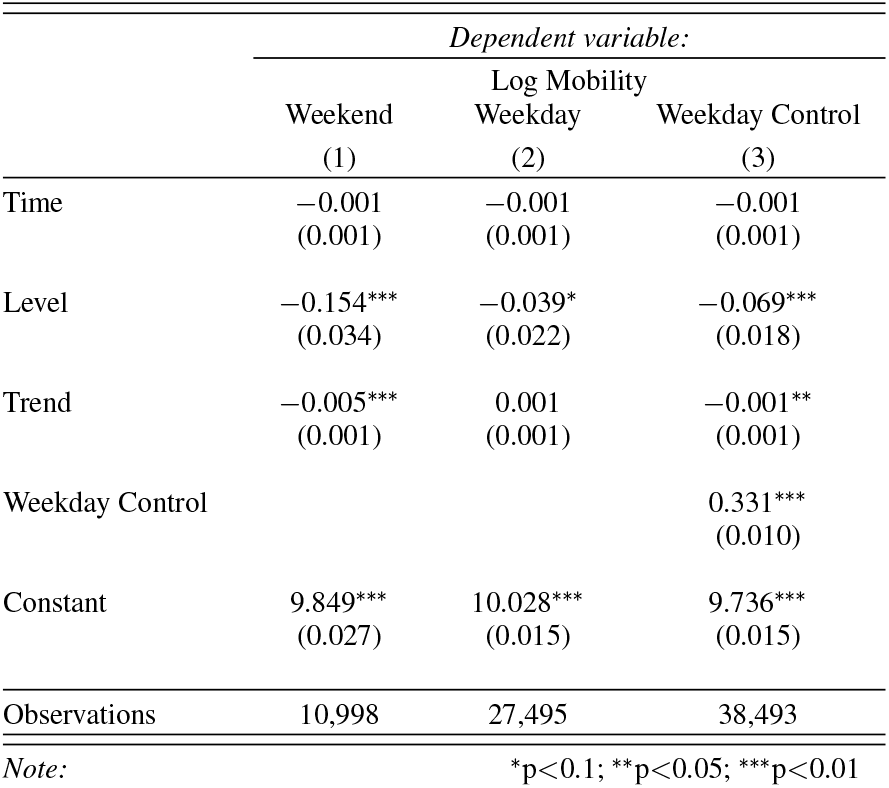
OLS Regression Results: The *Weekend* regression is only run on the data points on the weekend whereas the *Weekday* regression is only run on the data points on the weekdays. We introduce a weekend control, measuring both the weekends and weekdays mobility.

#### 2.2.2 Linear regression aggregated MITMA zones (Cataluña)

In order to visualise and illustrate the previous regression results in a more intuitive way, We aggregate the MITMA regions for all of Cataluña into daily mobility totals and thus we have a single observation for each day for Cataluña. Figure 4 plots the regression fitted values for the *incoming* mobility type before and after the policy with and without a weekday control variable. We note that the *level* variable of interest for Panel (A) is not statistically significant but the *level* variable in Panel (B) is statistically significant at the 5% level with a coefficient of -0.0840258 or -8.40%. Overall, both models show a drop in the regression line suggesting that mobility decreased after the policy.

**Figure 4.**
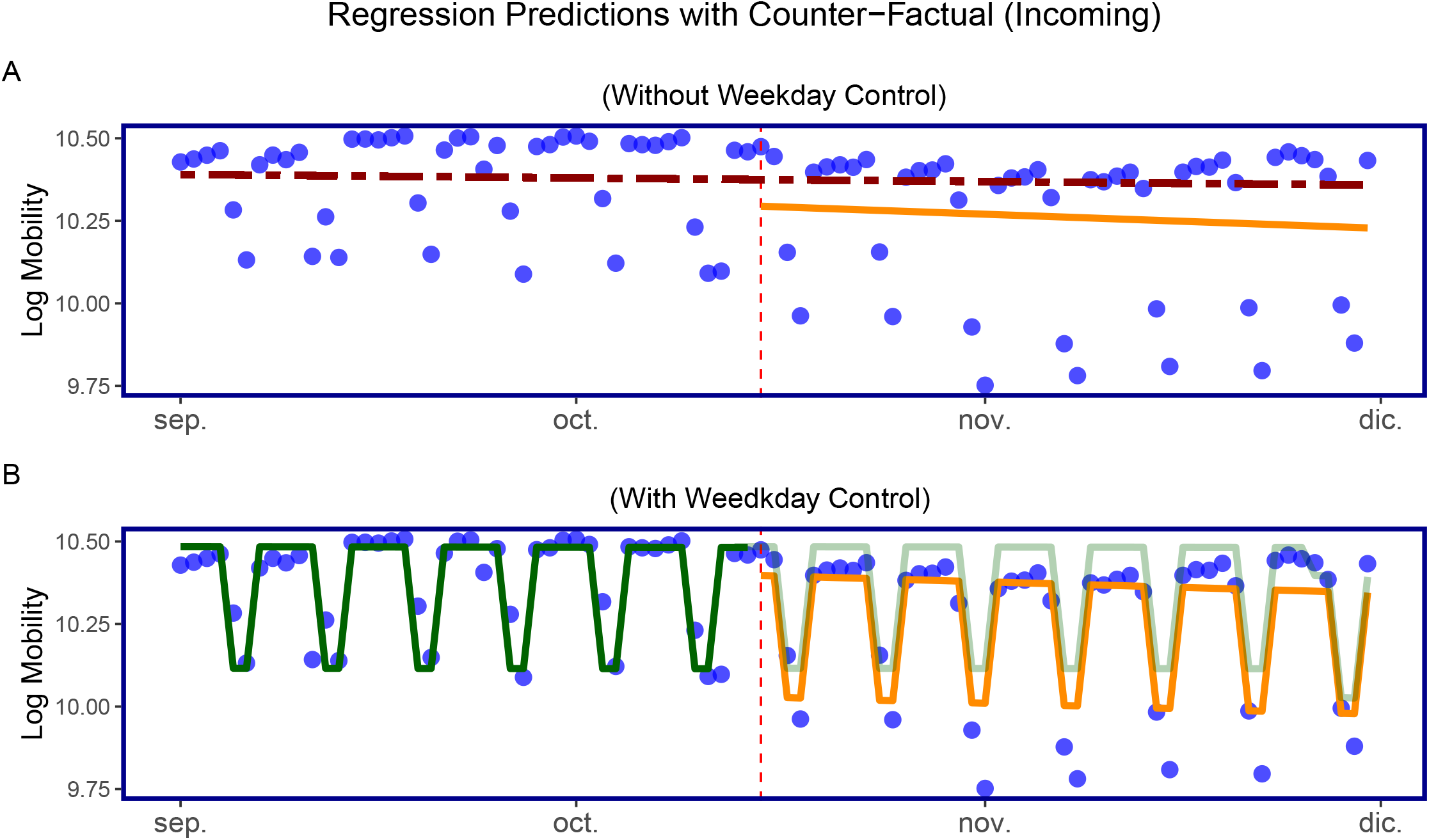
Linear regression model for total daily mobility in Cataluña: Panel (A) shows the fitted values from the regression without a weekday control whereas Panel (B) shows the fitted values with a weekday control. The vertical dotter line in both panels indicate the date in which the policy was introduced. The horizontal dotted line in Panel (A) after the policy shows the regression line had the policy not been implemented (fitted values from before the policy) whereas the orange line shows the fitted values of the regression after the policy. Panel (B) shows the fitted values before and after the’policy also in orange and the fitted values from before the policy in dark green and then lighter green after the policy.

### 2.3 Difference-in-differences

We next apply a difference-in-difference model in order to additionally quantify the effect on mobility of closing the bars and restaurant in Cataluña. Firstly, we use Madrid as a control group in order to visualise the difference-in-difference model since Madrid did not introduce the same policy of closing the bars and restaurants as in Cataluña, reported in Figure 5. Finally, we present the results using each autonomous community..

**Figure 5.**
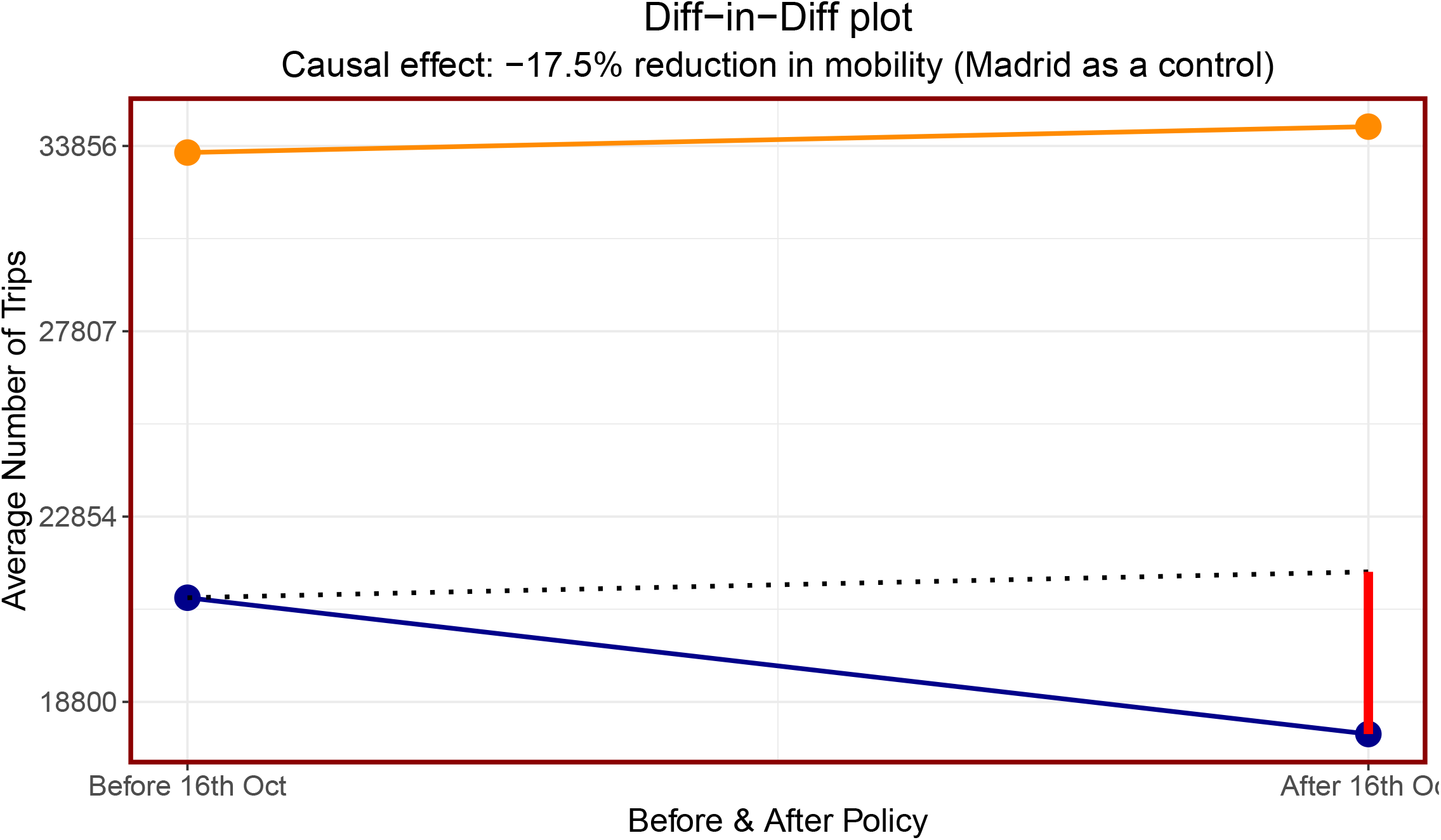
Difference-in-difference results for the *incoming* data: The 4 points correspond to the average number of trips before and after the policy was implemented, the orange colour corresponds to Madrid and the blue corresponds to Cataluña. The dotted line is Madrid’s line shifted downwards to show how potentially Cataluña’s mobility would have gone had they not implemented the policy. Finally, the vertical red line corresponds to the diff-in-diff regression coefficient as given in Equation 4. We find the causal effect from the model corresponds to a reduction in mobility of 17.5% when using Madrid as the control group. The y-axis has been re-scaled back to the number of trips.

We applied the difference-in-difference model to the four different mobility types *incoming, internal, outgoing* and *total* and extended the control groups to each autonomous community in Spain. (*Note: due to data handling issues, País Vasco and Castilla-La Mancha were omited*.) Table 2 reports the difference-in-difference estimators for each mobility type and autonomous community. Interestingly, the *internal* mobility does not show statistically significant results for all but three CCAAs (i.e. the cells in the table are not coloured). Recall, that *internal* mobility corresponds to the movements of people within a given MITMA zone. One economic interpretation could be that since bars and restaurants were closed people chose not to travel as much to different districts to socialise. For *internal* mobility, people still went about their daily business closer to their home, i.e. continue to shop at the supermarket and go to the pharmacy etc. but they had less incentive to travel to different districts (or MITMA regions) to meet with friends and family (which may be the reason for the non-statistically significant results in the *internal* mobility column). This result can be further seen in Figure 12 in the Appendix which plots a 14-day rolling moving average for 4 CCAA’s, the *internal* plot shows relatively flat lines for mobility, with Cataluña’s internal mobility only dropping slightly, whereas *incoming* and *outgoing* showed a much steeper drop for Cataluña relative to the other CCAA’s.

**Table 2.** Diff-in-Diff Estimates for Mobility Type: The mobility types *incoming* and *outgoing* appear correlated with all of the coefficients being similar to each other. Since these two mobility types are correlated, the *total* is also somewhat correlated. The CCAAs whose coefficients were statistically significant at the 0.1% level ranged between -9% to -17.5% and between -8.9% to -17.4% for *incoming* and *outgoing* respectively suggesting that the policy reduced mobility anywhere between -9% to -17.5% depending on the control group used. *Total* mobility at the same significance level ranged from -8.6% to -17.1%. These findings indicate that the policy had a real and direct effect on reducing the movement of people across MITMA regions.

| CCAA | Incoming | Internal | Outgoing | Total |
| --- | --- | --- | --- | --- |
| Andalucia | 1.2 | -3.1 | 1.3 | -0.7 |
| Aragon | -2.2 | -2.2 | -2 | -2.5 |
| Asturias | 1.7 | -0.2 | 1.7 | 0.7 |
| Balearsilles | -6.6 | -2.2 | -6.4 | -8 |
| Canarias | -12 | -5.6 | -11.9 | -12.4 |
| Cantabria | 1.4 | -0.6 | 1.3 | 0.8 |
| Castillaleon | -4.6 | 1.1 | -4.6 | -3.7 |
| Ceuta | -18 | -22.8 | -18 | -18.9 |
| Extremadura | -9 | -4.6 | -8.9 | -8.6 |
| Galicia | -2.6 | -3.1 | -2.5 | -3.3 |
| Larioja | -7.1 | -4.4 | -7 | -7.1 |
| Madrid | -17.5 | -8.3 | -17.4 | -17.1 |
| Melilla | -4.5 | 10.4 | -4.4 | -3.7 |
| Murcia | -4.1 | -4.7 | -4 | -6.2 |
| Navarra | -10.5 | -4.4 | -10.6 | -9.6 |
| Valencia | -11.7 | -7.4 | -11.7 | -11.4 |

Note:
All values in %
<sup>1</sup> 0.1% 0.05% 0.01% 0.001% significance levels

Therefore, people still moved within their MITMA regions in order to go about their daily lives but after the policy people stopped migrating to other regions and therefore longer distance mobility habits changed.

### 2.4 Bayesian structural time-series

Figure 6 shows the Bayesian structural time-series model in which the model uses Madrid’s mobility data in order to build a predictive model for Cataluña’s mobility. The model is able to build a very predictive model during the training phase. The large spike downwards in the *pointwise* panel, at the date 2020-09-11 in the training phase corresponds to *La Diada Nacional de Catalunya* a public holiday specific only to Cataluña and thus affects Cataluña mobility and not Madrid (Figure 3 shows that on public holidays mobility is reduced.). Aside from this single day the differences between the predictive model and the observed data lie in and around zero before the policy is implemented. After the introduction of the policy, these differences become negative except on two dates 2020-11-02 and 2020-11-09 which correspond to *Día de todos los Santos* and *Virgen de la Almudena* two public holidays specific to Madrid and not Cataluña.

**Figure 6.**
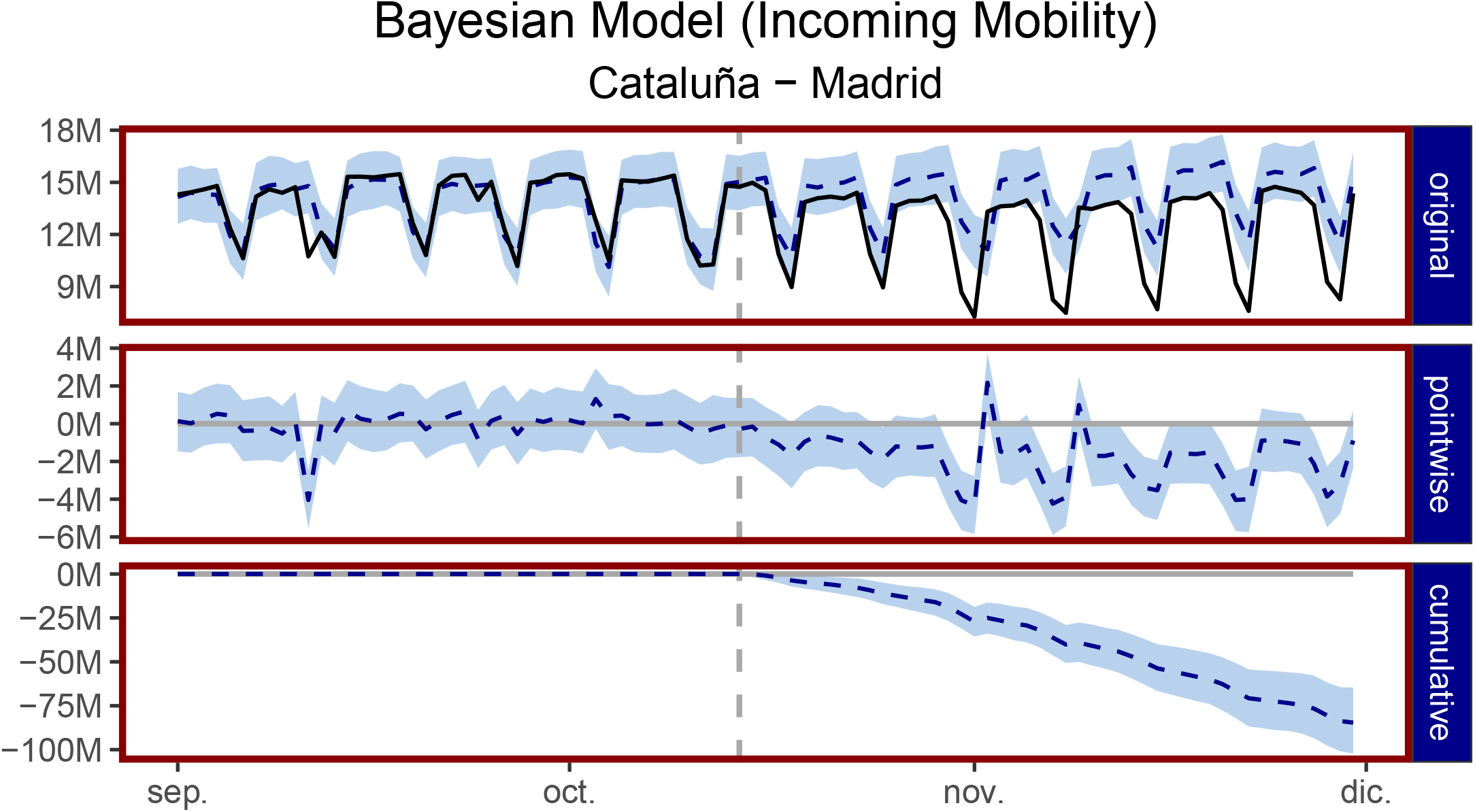
Bayesian structural time-series model: The *original* panel corresponds to the actual data and a counter-factual prediction for the post-treatment period. The *pointwise* panel shows the *pointwise* causal effect and it is the difference between the observed data and the counter-factual predictions. The *cumulative* panel shows the cumulative effect of the policy impact. Data before the dotted vertical line correspond to the observed data and a pre-policy predictive model, the data after the vertical dotted line corresponds to the observed data and a post-policy predictive model, that is, a synthetic counter-factual showing what the mobility in Cataluña may have looked like had the policy not been implemented.

We find that in absolute terms during the post-policy period, the average mobility in Cataluña was 12.39 million trips, in the absence of the policy the model expected an average of 14.19 million trips with a 95% confidence interval of (13.79, 14.58) million. The causal effect is 12.39 *−*14.19 = *−*1.8 million reduction in the number of trips in Cataluña with a 95% confidence interval (−2.19, -1.39) million. In relative terms, mobility reduced by -13% with a 95% confidence interval (−15%, -10%) which suggests that this causal effect in the reduction in mobility is statistically significant with a posterior Bayesian one-sided tail-area probability of 0.001. This finding is consistent with the linear regression and difference-in-difference models presented earlier. Figure 6 reports the model for Cataluña and Madrid, Figure 13 in the Appendix reports a case where the model did not work using Andalucia in place of Madrid as a control group. Finally, Table 3 shows the results for all CCAAs for the *incoming* mobility type, the *internal, outgoing* and *total* are left to the appendix.

**Table 3.** Bayesian Structural Time Series Estimates: The table can be analysed (using Madrid) that the reduction in trips in Cataluña fell by 12.7% with a confidence interval of (−15.7%, -9.9%) which is statistically significant at the 0.1% level. These findings are consistent with the results found in the difference-in-difference *internal* column of Table 2.

| CCAA | Incoming |  |  |  | P-value |
| --- | --- | --- | --- | --- | --- |
|  | Lower | Average | Upper | SD |  |
| Andalucia | -3.6 | 0.1 | 3.5 | 1.8 |  |
| Aragon | -9.3 | -6.3 | -3.3 | 1.6 | *** |
| Asturias | -5.3 | -1.5 | 2.5 | 2.0 |  |
| Balearsilles | -10.8 | -7.4 | -4.0 | 1.7 | *** |
| Canarias | -13.4 | -10.7 | -7.9 | 1.5 | *** |
| Cantabria | -7.1 | -3.2 | 0.8 | 2.0 | . |
| Castillaleon | -9.1 | -5.8 | -2.4 | 1.6 | *** |
| Ceuta | -16.3 | -13.3 | -10.2 | 1.5 | *** |
| Extremadura | -12.1 | -8.6 | -5.1 | 1.8 | *** |
| Galicia | -6.1 | -2.5 | 1.0 | 1.8 | . |
| Larioja | -10.0 | -6.4 | -3.0 | 1.8 | ** |
| Madrid | -15.7 | -12.7 | -9.9 | 1.4 | *** |
| Melilla | -7.1 | -3.2 | 0.8 | 2.1 | . |
| Murcia | -8.8 | -5.7 | -2.7 | 1.6 | *** |
| Navarra | -9.8 | -6.9 | -3.9 | 1.6 | ** |
| Valencia | -12.7 | -9.5 | -6.2 | 1.7 | *** |

Note:
All values in %
<sup>1</sup> \*\*\* 0.1%, \*\* 1%, \* 5%, . 10% significance levels

### 2.5 Mobility and COVID19

Thus far, this paper has shown that the policy has had a clear effect on the reduction of mobility in Cataluña. In this section we aim to assess the policies effect on daily COVID19 incidence, however, some comments are required.

Quantifying the policies impact on reductions in the number of new cases is more difficult than quantifying its impact on mobility. That is, mobility levels over time are roughly constant and predictable which allows us to build a strong counter-factual and thus when a policy shock has been implemented we can use this counter-factual in order to measure the real causal impact of that policy. Using new COVID19 case data presents more difficult problems since outbreaks are unpredictable and different regions experience fluctuations and peaks in COVID19 cases at different times, this renders the models used in this paper obsolete when applied to COVID19 case data. Consider Figure 1 which plots the number of new cases over time since the start of the pandemic. There are three regions with high peaks, Madrid experienced its peak before that of Cataluña and would therefore not make a good counter-factual after the policy. Andalucía and Cataluña peaked during the same period however, both CCAAs introduced strict counter measures to reduce the number of cases and both their number of new cases fell at the same time and therefore Andalucía would not make a suitable counter-factual either.

We compute the growth rate ratio in the number of cases for each CCAA as follows.

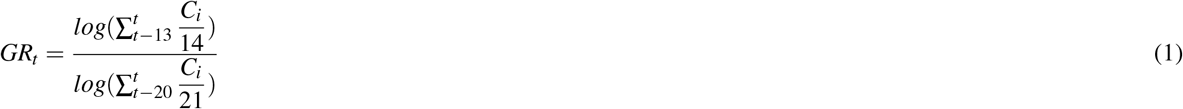

Where *C*_*i*_ is the number of cases for an autonomous community *i* at time *t*.

Figure 7 shows the growth rate ratio for the number of cases over time for all autonomous communities. The growth rate began to increase in the month leading up to when regional governments introduced different policies. The growth rate in Cataluña began to decrease after an initial 2-3 week lag once the policy came into effect. Figure 14 shows that the growth rate Figure 8 shows the same calculation as in equation 1 but applied to the mobility time series. The growth rate ratio for mobility began to increase from September for many CCAAs, this could be attributed to people returning from their second homes after the summer break.

**Figure 7.**
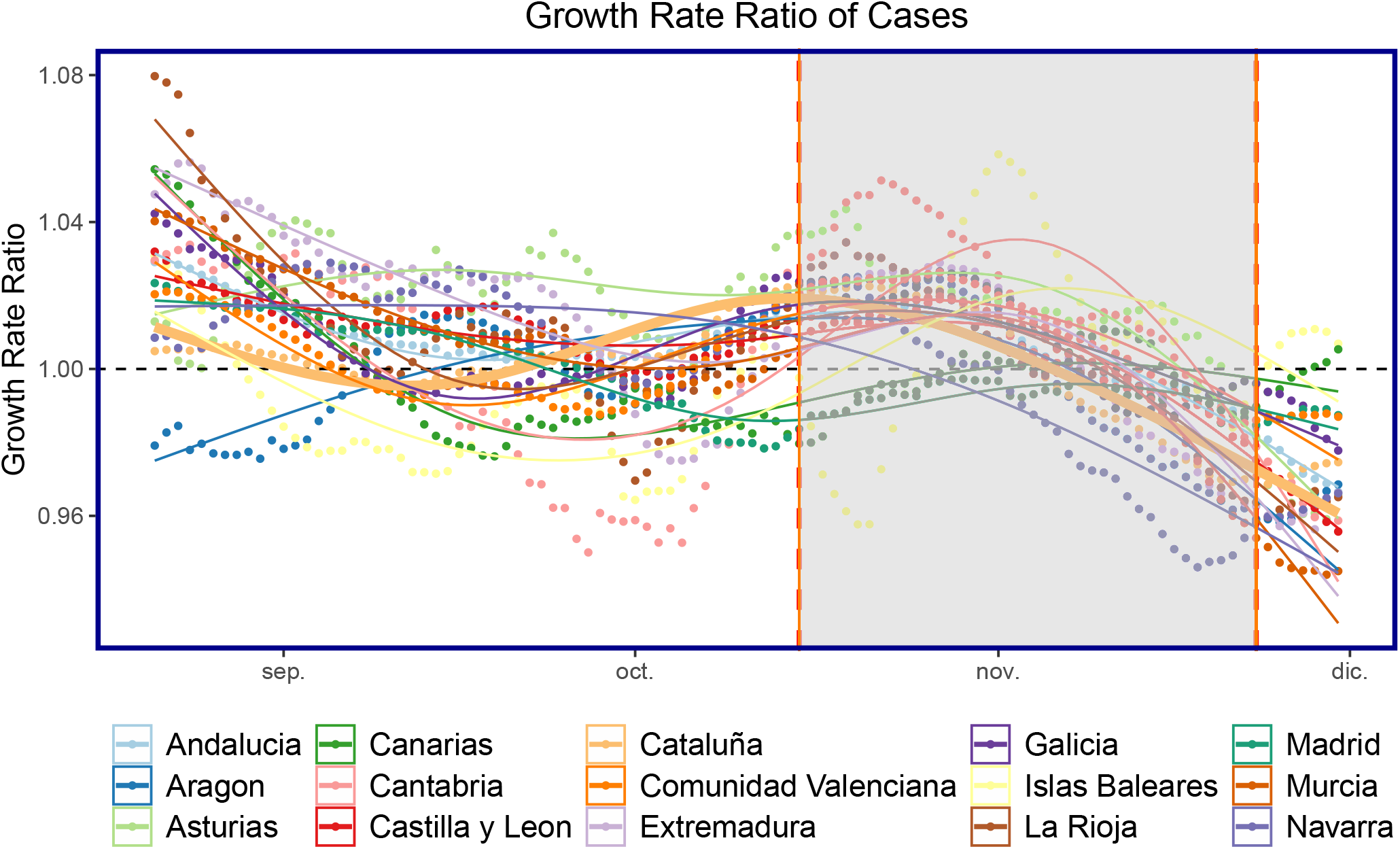
Growth Rate Ratio (GR) of cases: All autonomous communities are shown as points with Cataluña shown as the thicker line plot for ease of composition. The shaded area corresponds to when the policy of closing bars and restaurants in Cataluña was implemented. in the number of cases began to increase in December, before Christmas and when the policy was not enforced.

**Figure 8.**
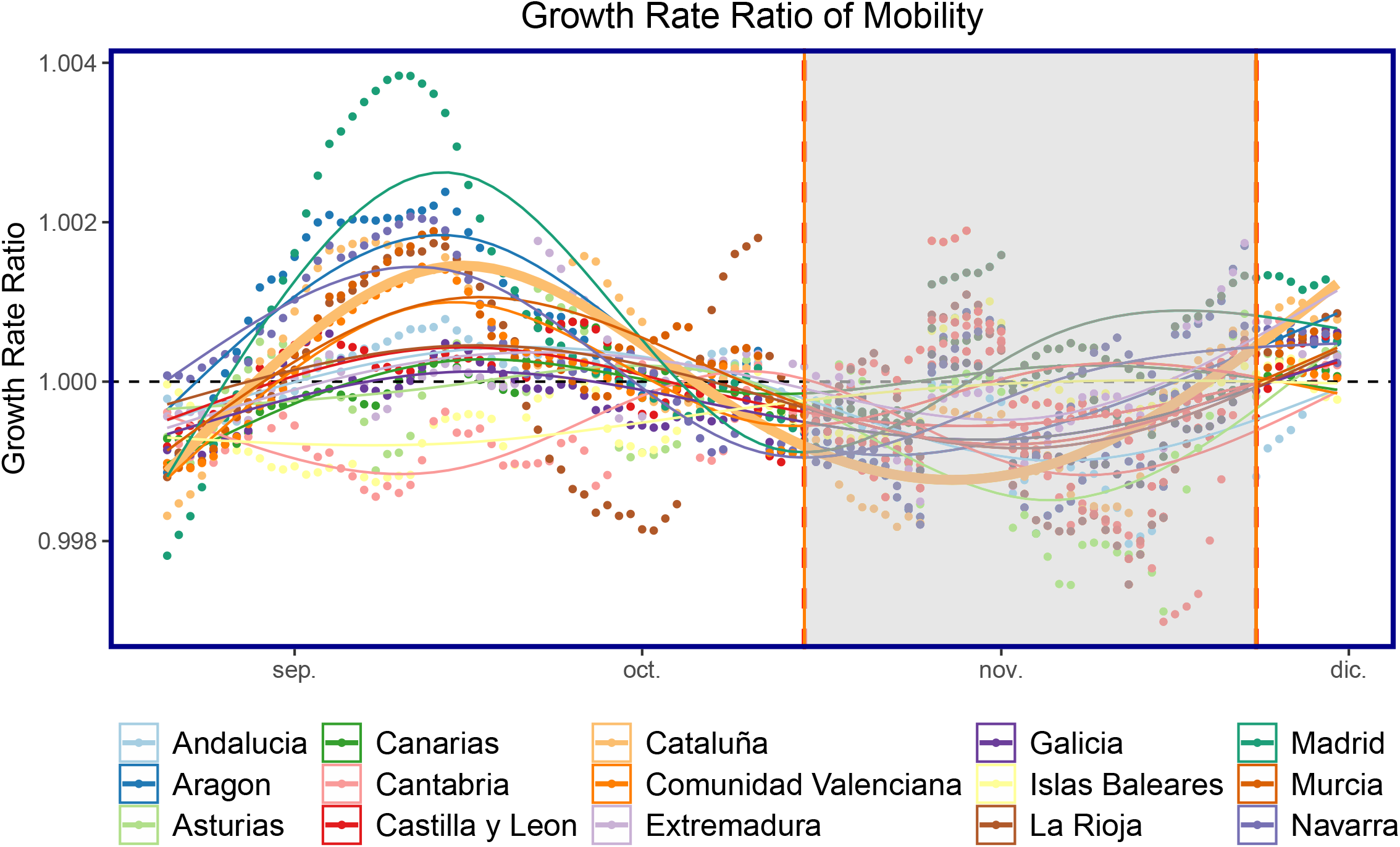
Growth Rate Ratio (GR) of mobility: All autonomous communities are shown as points with Cataluña shown as the thicker line plot for ease of composition. The shaded area corresponds to when the policy of closing bars and restaurants in Cataluña was implemented.

We next anticipate that there is a lag between the movement of people and becoming infected with COVID19, due to the lag in symptoms, reporting etc. We firstly normalise the mobility data, fixed to the first week of October as follows in equation 2. The first week of October is the first week in our sample data. Moreover, the data is normalised such that, each Monday is normalised to the first Monday of October, each Tuesday is normalised to the first Tuesday of October and so on.

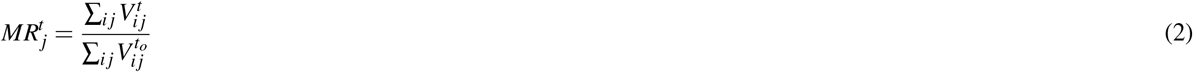

Where *t*_*o*_ corresponds to the mobility data for each corresponding weekday at the beginning of October. Next, we compute the Pearson correlation coefficients with a 95% confidence interval for lags 1:40 as shown in Figure 9. We finally take the optimal lag of 21 and plot the scatter-plot between the growth rate ratio defined in equation 1 and the normalised mobility defined in equation 2. The results can be seen in Figure 10. There is an association between increased mobility and an increase in the growth rate ratio.

**Figure 9.**
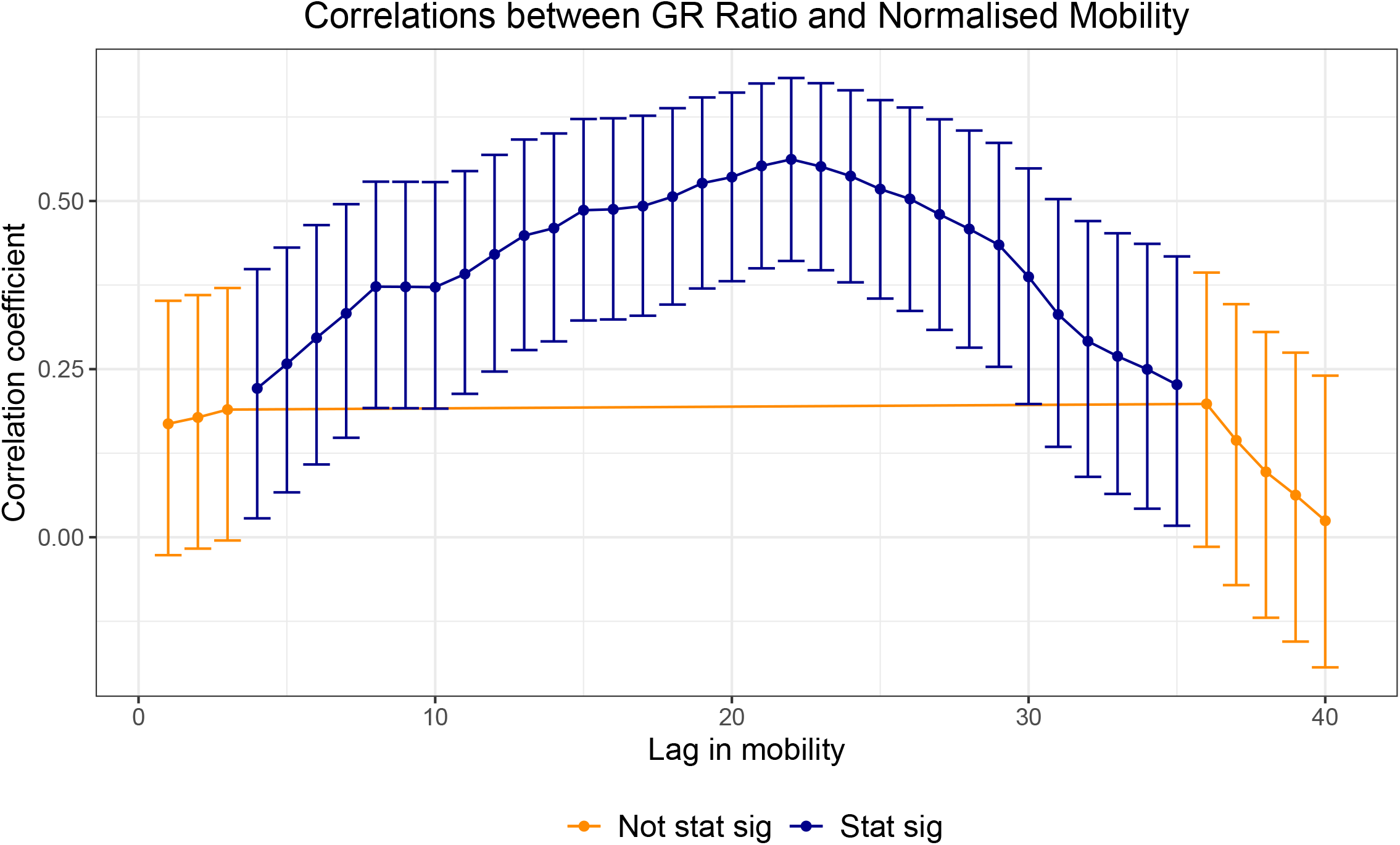
Normalised correlations: Relationship between the growth rate ratio and mobility-normalised. The optimal lag occurs around 21 days. This may account for lags in the time people notice symptoms of COVID19 after some days and lags in the reporting of cases for each regional healthcare systems.

**Figure 10.**
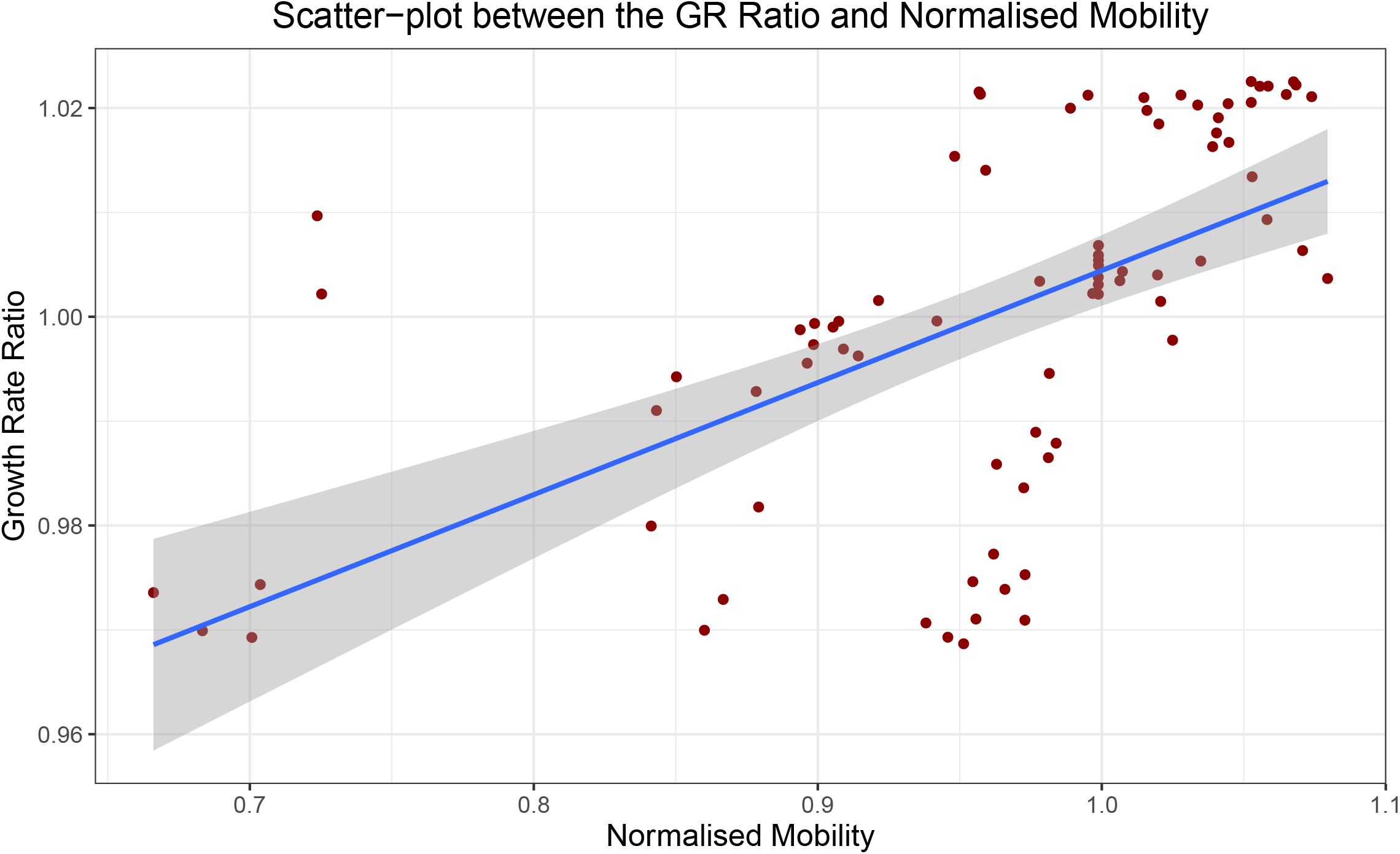
Scatter-plot between the Growth Rate Ratio in the number of cases and the normalised mobility: There appears to be some relationship between an increase in mobility and an increase in the growth rate ratio for the optimal lag of 21 days.

## 3 Conclusion

This paper quantifies the impact of mobility on the closing of bars and restaurants in Cataluña using a number of causal inference models. Overall we find that this policy reduced mobility in Cataluña, but it not only reduced mobility, it caused people to change the way they behave and respond to the policy, with trips across regions being affected more than trips within regions. This finding is significant since people did not substitute meeting friends and family at bars and restaurants with meeting friends and family in other locations, people simply stayed within their own MITMA region, reducing their overall mobility and changed the way in which people went about their daily business. The findings of this paper are 3-fold. (a) we quantify the impact of the policy of closing bars and restaurants down during a 5 week period on mobility. (b) we find that people changed their social behaviours in a direct response to the policy, reducing their mobility relatively more so on weekends than on weekdays, additionally people changed their longer distance travel habits whilst leaving shorter distance travel habits relatively unchanged. (c) we find that there is some evidence that the policy slowed the growth rate of COVID19 incidence in Cataluña.

## 4 Methods

### 4.1 Mobility Data Records

Mobility data records comes from a study conducted by the Ministerio de Transportes, Movilidad y Agenda Urbana (MITMA) https://www.mitma.gob.es/ministerio/covid-19/evolucion-movilidad-big-data. The study collects data of mobility and distribution of the population in Spain from 13 million anonymised mobile-phone lines provided by a single mobile operator whose subscribers are evenly distributed. More specifically, the data is reported on a geographical layer composed of 2850 mobility zones across the whole of Spain where each mobility zone corresponds to a district or group of districts in densely populated areas, and to municipalities or groups of municipalities in regions with low population density (see Figure 11). The unit to measure mobility is the trip and the data contains the number of trips between and within mobility zones reported in and hourly basis. The start of a trip event is defined as when a user moves more than 500 meters and the end of the trip is defined as when that user remains in an antenna coverage area for more than 20 minutes. Thus, a person moving from region *𝒜* to region *ℬ* and remains in region *ℬ* for longer than 20 minutes is defined as a trip *between* regions. Moreover a person who remains in region *𝒜* and is connected to antenna *𝒜*_*i*_ then moves to antenna *𝒜*_*j*_ is defined as a trip *within* region. We collected mobility data from 01-09-2020 to 30-11-2020 for all 17 autonomous communities in Spain using the Flow-Maps systems. https://github.com/bsc-flowmaps/ *Note: due to data handling issues, País Vasco and Castilla-La Mancha were omited*. Figure 11 shows the different mobility zones for all of Spain, the colours do not hold meaning and are simply coloured for easier illustration of the different MITMA areas.

**Figure 11.**
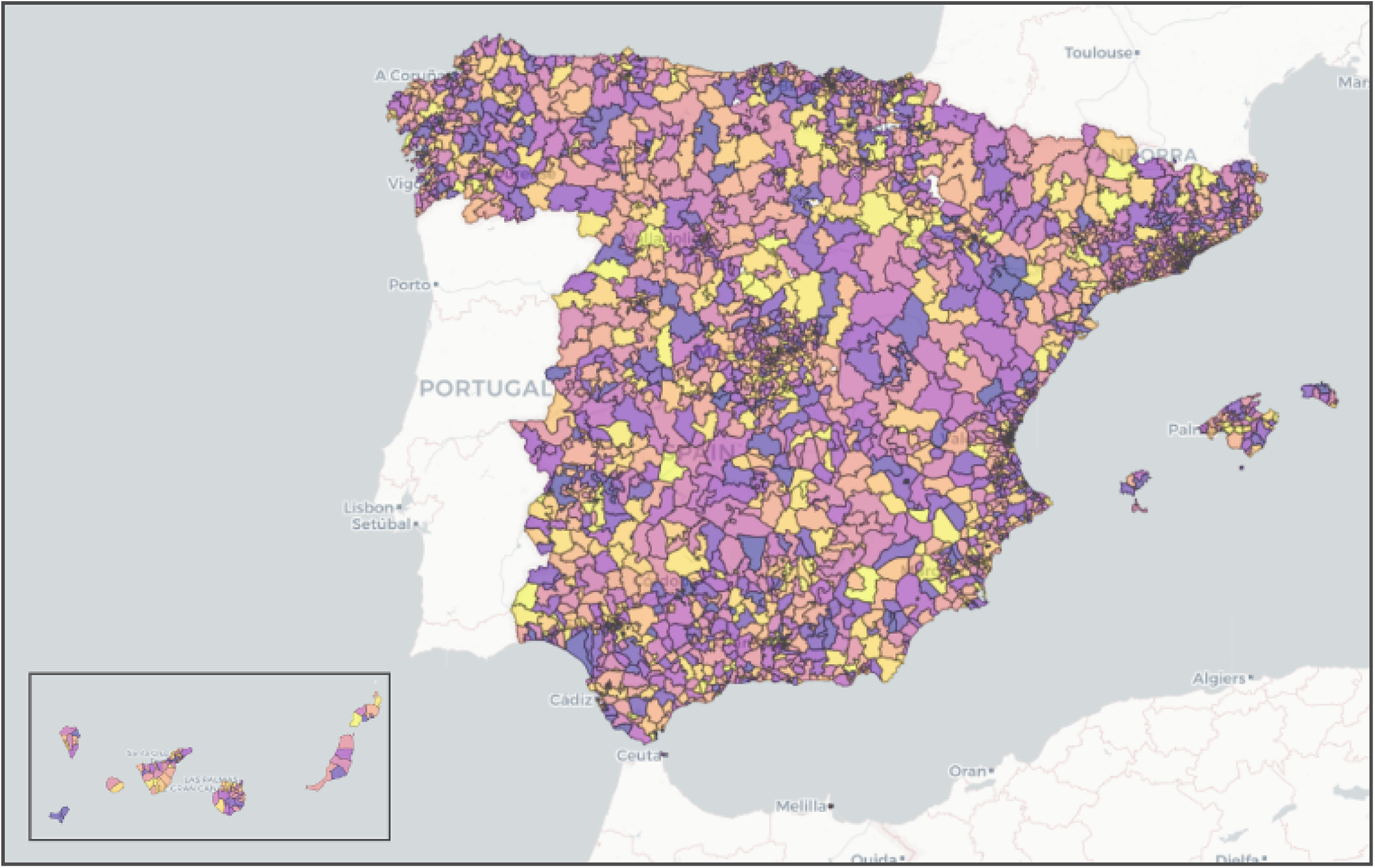
MITMA Regions: Each coloured polygon corresponds to a mobility zone. Naturally, smaller mobility zones are located in densely populated areas such as Madrid, Barcelona and along the coast.

**Figure 12.**
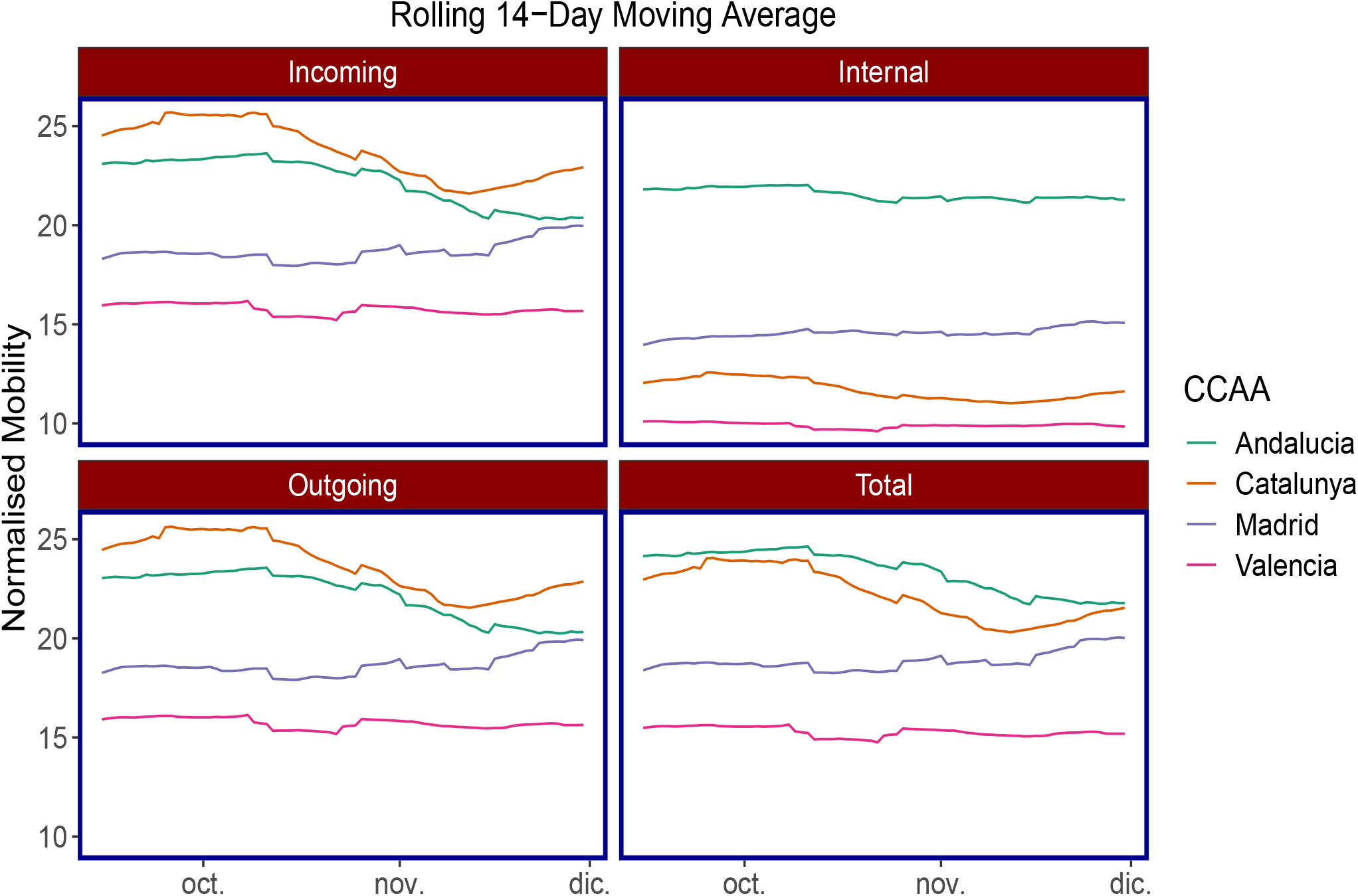
14-Day rolling average for different mobility types. Only the top 4 CCAA’s are reported for ease of composition. It is evident that after the policy, Cataluña’s mobility dropped significantly for *incoming, outgoing* and *total* and began increasing once the restrictions on bars and restaurants eased. The same can not be said for the *internal* mobility type.

**Figure 13.**
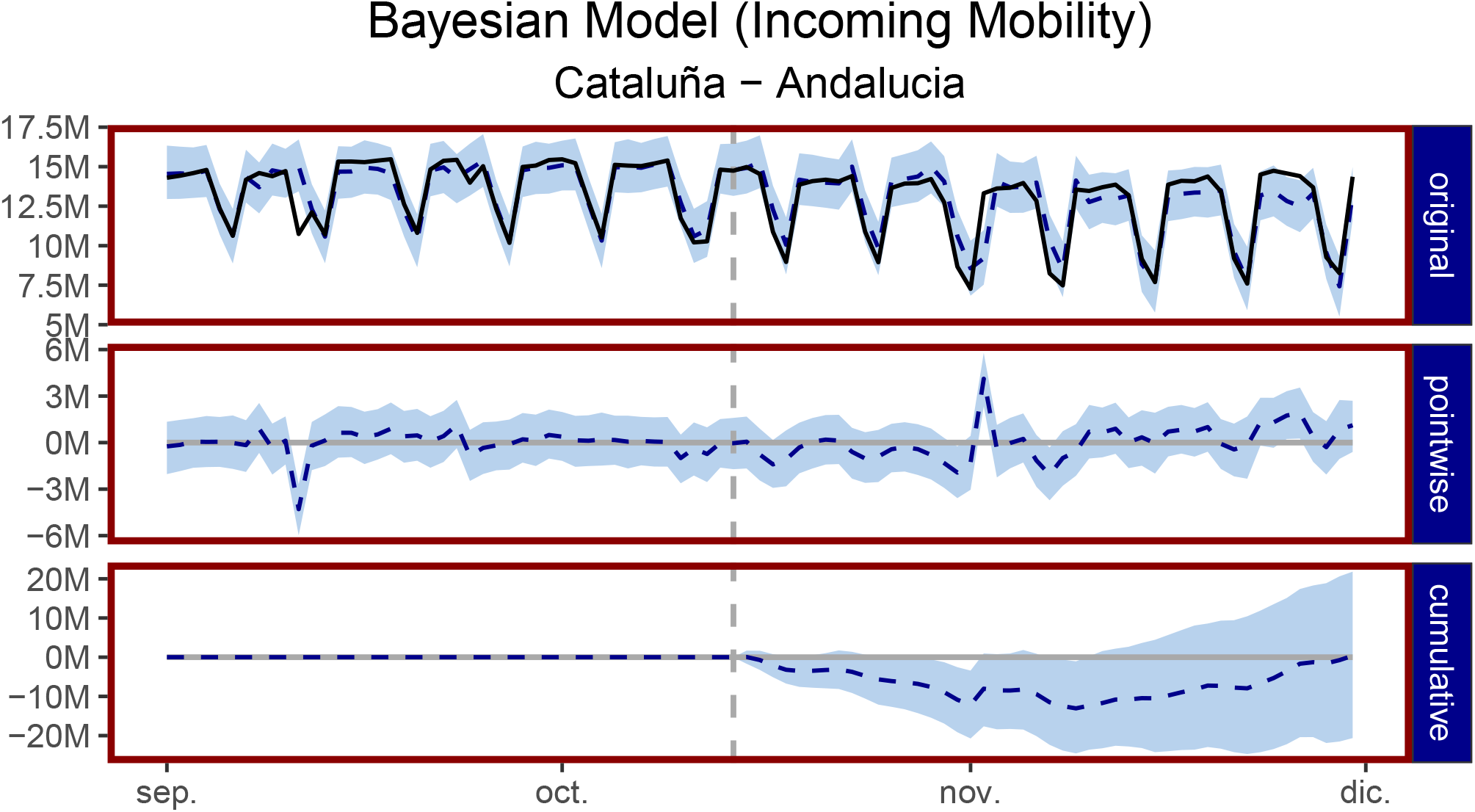
Bayesian structural time-series model: This figure follows on from Figure 6 in which we report a case where the control was not suitable for Cataluña. The pairwise differences hover around zero and the cumulative mobility returns back to zero suggesting that causal inference cannot be inferred from this control group. As Table 3 shows, Andalucia is not statistically significant and the sign of the relative effect is positive, not negative as one would expect.

**Figure 14.**
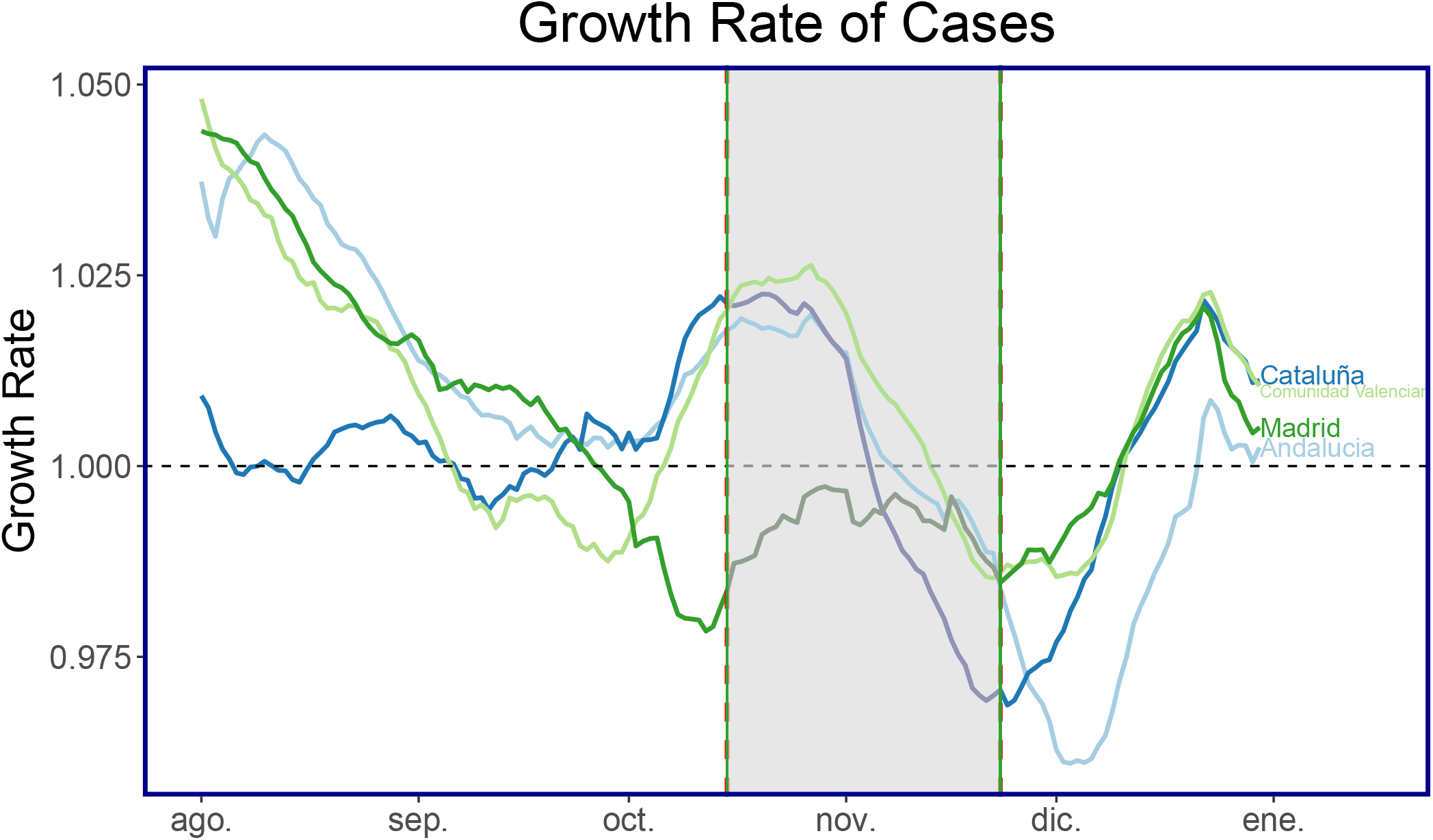
Growth Rate (GR) of cases from August until December for four of the most populous regions in Spain. The shaded area corresponds to when the policy of closing bars and restaurants was implemented.

After retrieving the raw mobility data reported by hour, we aggregate it by summing the the total trips between and within mobility zones for each individual day, to obtain mobility reported on a daily basis. Using the daily mobility data we construct *origin-destination* (OD) matrices for each day *t*. Table 4.1 represent and example of OD matrix where rows correspond to the *origin* zone and the columns correspond to the *destination* zone and each entry corresponds to the total daily trips from *origin* to *destination*.

|  |  | Destination<sub>t</sub> |  |  |  |
| --- | --- | --- | --- | --- | --- |
| | | $\mathcal{A}_d$ | $\mathcal{B}_d$ | ... | $\mathcal{Z}_d$ |
| Origin<sub>t</sub> | $\mathcal{A}_o$ | 5639 | 1873 | | 1072 |
| | $\mathcal{B}_o$ | 2192 | 7692 | | 1965 |
|  | ... |  |  | ... |  |
| | $\mathcal{Z}_o$ | 1129 | 1834 | | 6393 |

**Table 4.** Diff-in-diff: Difference-in-difference estimates

|  | Pre | Post | Post-Pre Difference |
| --- | --- | --- | --- |
| **Treatment** | $\bar{y}_0^T$ | $\bar{y}_1^T$ | $\bar{y}_1^T - \bar{y}_0^T$ |
| **Control** | $\bar{y}_0^C$ | $\bar{y}_1^C$ | $\bar{y}_1^C - \bar{y}_0^C$ |
| **T-C Difference** | $\bar{y}_0^T - \bar{y}_0^C$ | $\bar{y}_1^T - \bar{y}_1^C$ | $\bar{y}_1^T - \bar{y}_1^C - (\bar{y}_0^T - \bar{y}_0^C)$ |

**Table 5.**
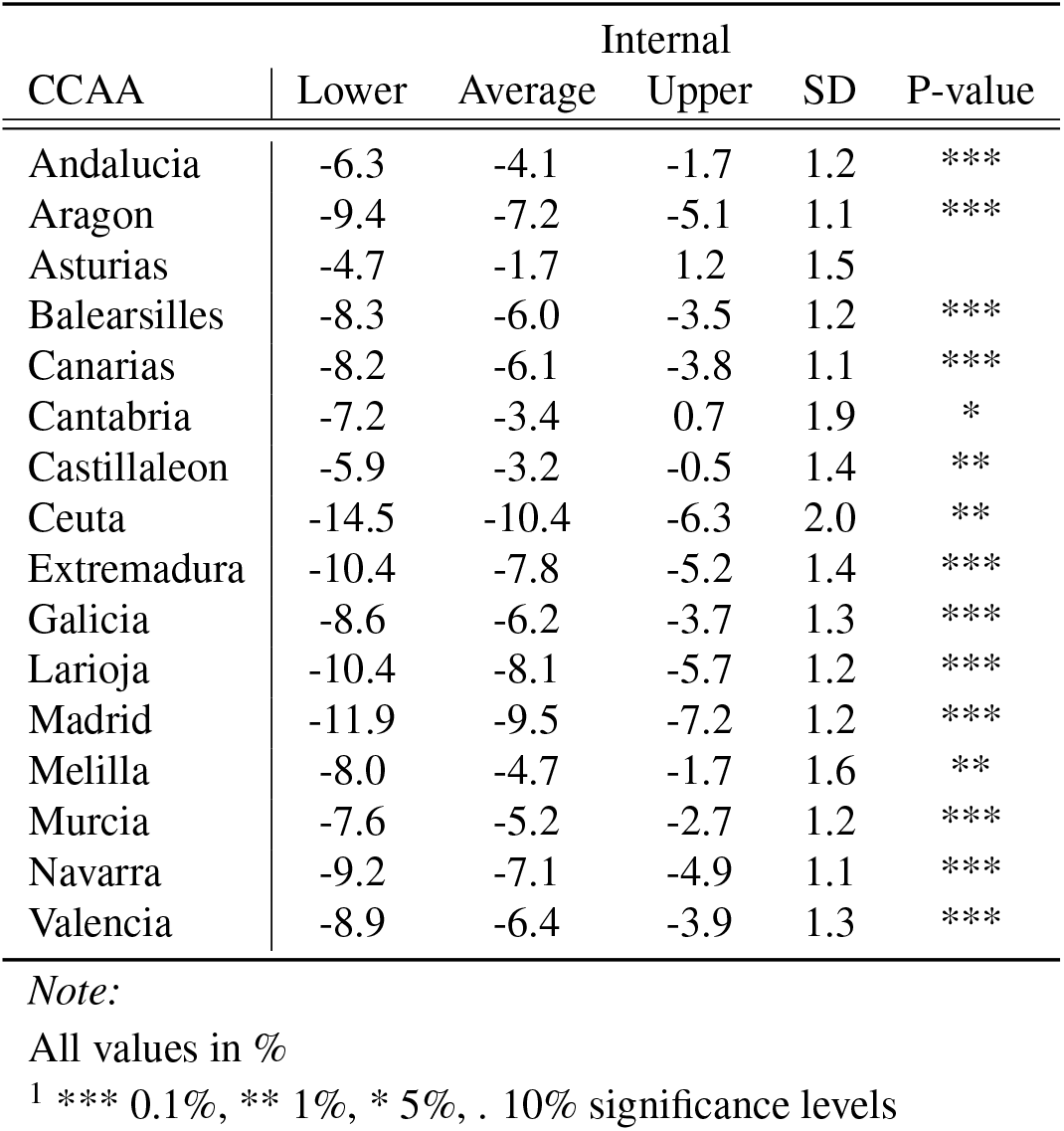
Bayesian Structural Time Series Estimates.

| CCAA | Internal |  |  |  | P-value |
| --- | --- | --- | --- | --- | --- |
|  | Lower | Average | Upper | SD |  |
| Andalucía | -6.3 | -4.1 | -1.7 | 1.2 | *** |
| Aragón | -9.4 | -7.2 | -5.1 | 1.1 | *** |
| Asturias | -4.7 | -1.7 | 1.2 | 1.5 |  |
| Baleares | -8.3 | -6.0 | -3.5 | 1.2 | *** |
| Canarias | -8.2 | -6.1 | -3.8 | 1.1 | *** |
| Cantabria | -7.2 | -3.4 | 0.7 | 1.9 | * |
| Castilla y León | -5.9 | -3.2 | -0.5 | 1.4 | ** |
| Ceuta | -14.5 | -10.4 | -6.3 | 2.0 | ** |
| Extremadura | -10.4 | -7.8 | -5.2 | 1.4 | *** |
| Galicia | -8.6 | -6.2 | -3.7 | 1.3 | *** |
| Larioja | -10.4 | -8.1 | -5.7 | 1.2 | *** |
| Madrid | -11.9 | -9.5 | -7.2 | 1.2 | *** |
| Melilla | -8.0 | -4.7 | -1.7 | 1.6 | ** |
| Murcia | -7.6 | -5.2 | -2.7 | 1.2 | *** |
| Navarra | -9.2 | -7.1 | -4.9 | 1.1 | *** |
| Valencia | -8.9 | -6.4 | -3.9 | 1.3 | *** |

*Note:*
All values in %
<sup>1</sup> \*\*\* 0.1%, \*\* 1%, \* 5%, . 10% significance levels

**Table 6.** Bayesian Structural Time Series Estimates.

| CCAA | Outgoing |  |  | SD | P-value |
| --- | --- | --- | --- | --- | --- |
|  | Lower | Average | Upper |  |  |
| Andalucía | -3.6 | 0.1 | 3.8 | 1.8 |  |
| Aragón | -9.4 | -6.4 | -3.3 | 1.6 | *** |
| Asturias | -5.3 | -1.5 | 2.5 | 2.0 |  |
| Baleares | -10.5 | -7.4 | -4.0 | 1.7 | *** |
| Canarias | -13.4 | -10.7 | -7.7 | 1.5 | *** |
| Cantabria | -6.9 | -3.2 | 0.8 | 1.9 | . |
| Castilla y León | -8.6 | -5.8 | -2.4 | 1.6 | *** |
| Ceuta | -16.2 | -13.3 | -10.4 | 1.5 | *** |
| Extremadura | -11.9 | -8.6 | -5.2 | 1.7 | *** |
| Galicia | -6.2 | -2.5 | 1.1 | 1.8 | . |
| Larioja | -9.8 | -6.4 | -2.8 | 1.8 | *** |
| Madrid | -15.4 | -12.7 | -9.8 | 1.4 | *** |
| Melilla | -7.3 | -3.2 | 0.9 | 2.1 | . |
| Murcia | -8.8 | -5.7 | -2.5 | 1.6 | ** |
| Navarra | -9.9 | -7.0 | -3.8 | 1.5 | *** |
| Valencia | -12.7 | -9.5 | -6.1 | 1.7 | *** |

*Note:*
All values in %
<sup>1</sup> \*\*\* 0.1%, \*\* 1%, \* 5%, . 10% significance levels

**Table 7.**
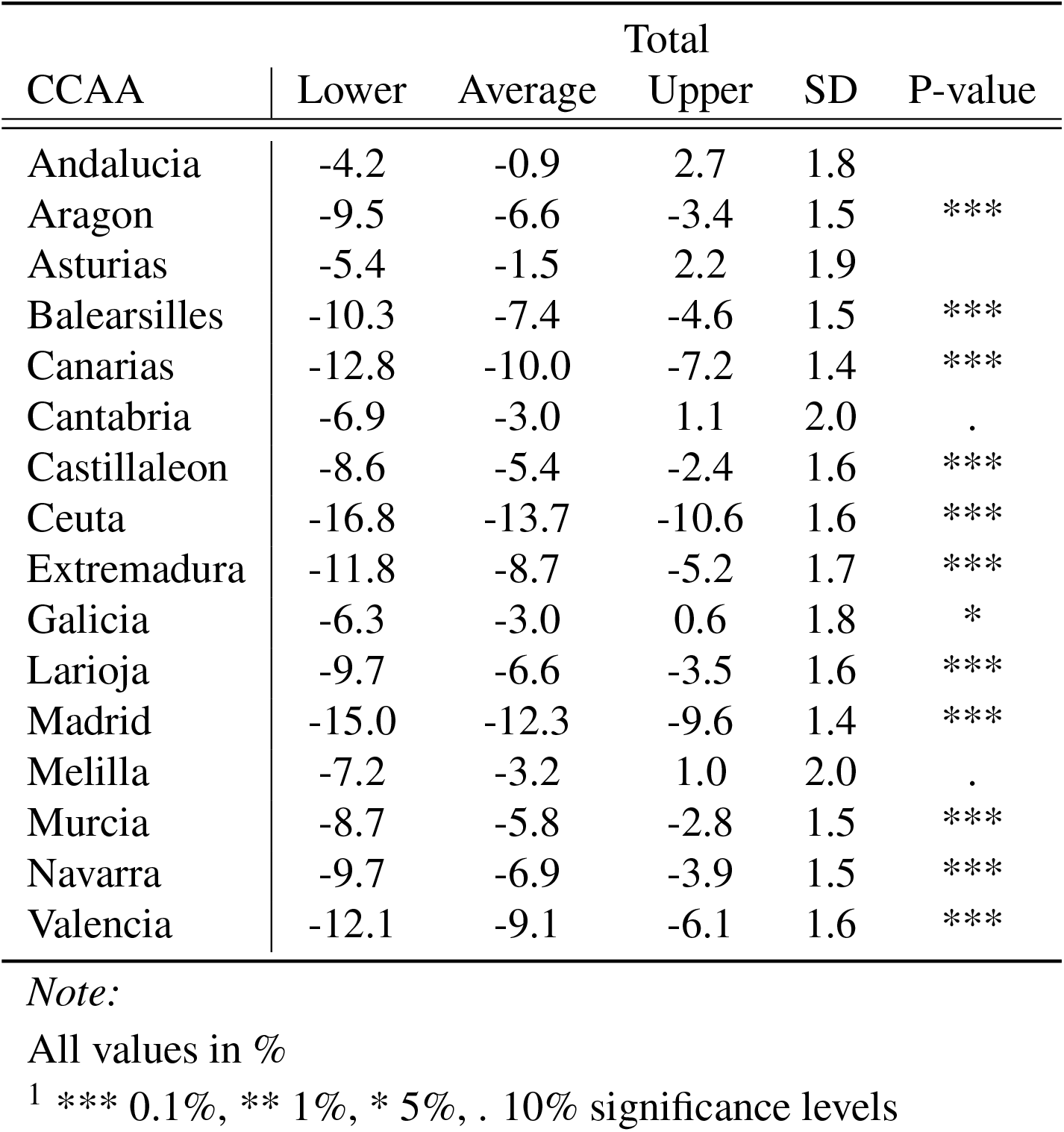
Bayesian Structural Time Series Estimates.

| CCAA | Total |  |  |  | P-value |
| --- | --- | --- | --- | --- | --- |
|  | Lower | Average | Upper | SD |  |
| Andalucia | -4.2 | -0.9 | 2.7 | 1.8 |  |
| Aragon | -9.5 | -6.6 | -3.4 | 1.5 | *** |
| Asturias | -5.4 | -1.5 | 2.2 | 1.9 |  |
| Balearsilles | -10.3 | -7.4 | -4.6 | 1.5 | *** |
| Canarias | -12.8 | -10.0 | -7.2 | 1.4 | *** |
| Cantabria | -6.9 | -3.0 | 1.1 | 2.0 | . |
| Castillaleon | -8.6 | -5.4 | -2.4 | 1.6 | *** |
| Ceuta | -16.8 | -13.7 | -10.6 | 1.6 | *** |
| Extremadura | -11.8 | -8.7 | -5.2 | 1.7 | *** |
| Galicia | -6.3 | -3.0 | 0.6 | 1.8 | * |
| Larioja | -9.7 | -6.6 | -3.5 | 1.6 | *** |
| Madrid | -15.0 | -12.3 | -9.6 | 1.4 | *** |
| Melilla | -7.2 | -3.2 | 1.0 | 2.0 | . |
| Murcia | -8.7 | -5.8 | -2.8 | 1.5 | *** |
| Navarra | -9.7 | -6.9 | -3.9 | 1.5 | *** |
| Valencia | -12.1 | -9.1 | -6.1 | 1.6 | *** |

*Note:*
All values in %
<sup>1</sup> \*\*\* 0.1%, \*\* 1%, \* 5%, . 10% significance levels

Following the example of Table 4.1, trips from *𝒜*_*o*_ to *ℬ*_*d*_ are associated with trips leaving MITMA zone *𝒜*_*o*_ and going to MITMA zone *ℬ*_*d*_ and are the outgoing trips from MITMA zone *𝒜*_*o*_. Incoming trips are defined as the opposite and thus trips which began at *ℬ*_*d*_ and went to *𝒜*_*o*_. The trips going from *𝒜*_*o*_ to *𝒜*_*d*_ are internal trips (the diagonal of the matrix) or the number of trips within MITMA zone *𝒜*. Furthermore, for each mobility zone on a given date we define the following four mobility indexes: i) *incoming* mobility; ii) *outgoing* mobility; iii) *internal* mobility; and iv) the *total* mobility. More formally, the incoming data takes the column sums of the OD matrix minus the diagonal, the outgoing data takes the row sums of the OD matrix minus the diagonal, the internal is just the diagonal and the total is incoming + outgoing + internal.

### 4.2 Linear regression

We first run a linear regression model in order to see the effects on weekday and weekend mobility. We expect to see that the closure of bars and restaurants will have a greater effect on the weekend then on the weekdays since mobility is largely unaffected by people commuting to and from work but is affected by peoples decision to go and socialise on the weekends. We estimate the following for policy status *j*, at time *t*:

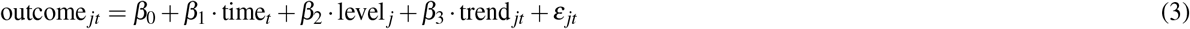

Where, outcome_*jt*_ is mobility for MITMA region *j* at time *t, β*_0_ is the intercept of the existing level at point 0, *β*_1_ gives the existing trend in mobility before the policy, *level* takes on a value of 0 before the policy and 1 after the policy with its *β*_2_ capturing the impact of the policy, *β*_3_ captures the change in trend.

### 4.3 Difference-in-differences (DiD)

We use a **Difference-in-difference** technique to infer the causal impact of a policy. That is, with two groups and two periods, the DiD estimator is defined as the difference in average outcome in the treatment group before and after treatment *minus* the difference in average outcome in the control group before and after treatment. Here the outcome variable is a daily time-series of mobility data and therefore we are considering mobility before the policy intervention and mobility after the policy intervention. Consider equation 4 in which Cataluña corresponds to the treatment group, denoted as *T* and other CCAAs, for instance Madrid corresponds to the control group, denoted as *C*. Therefore, 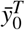corresponds to the average mobility for a given mobility zone in Cataluña before the policy, 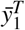 and the average mobility in a given mobility zone in Cataluña after the policy and 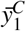 and 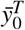 corresponds to the average mobility in a given mobility zone in a CCAA (excluding Cataluña) before and after the policy. Thus, the causal impact of the policy can be given as the difference of these two differences. It is important to note that the data is normalised 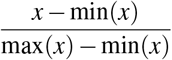, bounded by [0, 1].

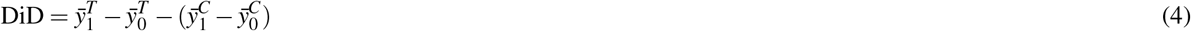

### 4.4 Bayesian structural time-series

We use **Bayesian structural time series** to infer the causal impact of a policy by explicitly modelling the counter-factual observed before and after an intervention (see (29) and (30)). The model assumes that the outcome time series can be explained in terms of a set of control time series that were themselves not affected by the intervention. Furthermore, the relation between the treated series and control series is assumed to be stable during the post-intervention period. This allow us to generalises the results obtained using the difference-in-difference approach. The model is first estimated using pre-intervention data, and then an intervention period occurs and the model then tries to predicts the post-intervention period. The difference between the prediction and the observed data can be thought of as the causal impact of the policy. For the pre-intervention period we take mobility before the policy and for the post-intervention period we take mobility after the policy.

Consider the following state-space model for time-series data.

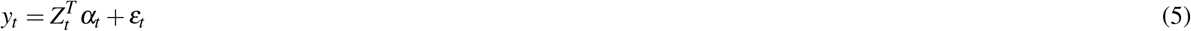

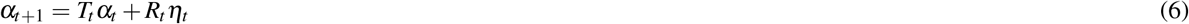

In which,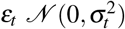 and *η*_*t*_ *𝒩* (0, *Q*_*t*_) are independent of all other unknowns. The *observation equation* 5 *links the observed data y*_*t*_ to a latent *d*-dimensional state vector *α*_*t*_ and the *state equation* 6 governs the evolution of the state vector *α*_*t*_ through time. *y*_*t*_ is a scalar, *Z*_*t*_ is a *d*-dimensional output vector, *T*_*t*_ is a *d ×d* transition matrix, *R*_*t*_ is a *d ×q* control matrix, *ε*_*t*_ is a scalar observation error with noise variance *σ*_*t*_ and *η*_*t*_ is a *q*-dimensional system error with a *q× q* state-diffusion matrix *Q*_*t*_, where *qd*. More details can be found in (30).

## Data Availability

All data is publically available via the FlowMaps website and Python package: https://flowmaps.life.bsc.es/flowboard/

https://pypi.org/project/flowmaps-data/

https://github.com/bsc-flowmaps

https://www.nature.com/articles/s41597-021-01093-5

## 5 Appendix

## Notes

## References

1. Liu, Y., Morgenstern, C., Kelly, J., Lowe, R. & Jit, M. The impact of non-pharmaceutical interventions on sars-cov-2 transmission across 130 countries and territories. BMC medicine 19, 1–12 (2021).

2. Hale, T., Petherick, A., Phillips, T. & Webster, S. Variation in government responses to covid-19. Blavatnik school government working paper 31, 2020–11 (2020).

3. Duhon, J., Bragazzi, N. & Kong, J. D. The impact of non-pharmaceutical interventions, demographic, social, and climatic factors on the initial growth rate of covid-19: A cross-country study. Sci. The Total. Environ. 760, 144325 (2021).

4. Aksoy, C. G., Ganslmeier, M. & Poutvaara, P. Public attention and policy responses to covid-19 pandemic. Available at SSRN 3638340 (2020).

5. Rocklöv, J. & Sjödin, H. High population densities catalyse the spread of covid-19. J. travel medicine 27, taaa038 (2020).

6. Bhadra, A., Mukherjee, A. & Sarkar, K. Impact of population density on covid-19 infected and mortality rate in india. Model. Earth Syst. Environ. 7, 623–629 (2021).

7. Kadi, N. & Khelfaoui, M. Population density, a factor in the spread of covid-19 in algeria: statistic study. Bull. Natl. Res. Centre 44, 1–7 (2020).

8. Rader, B. et al. Crowding and the shape of covid-19 epidemics. Nat. medicine 26, 1829–1834 (2020).

9. Hamidi, S., Sabouri, S. & Ewing, R. Does density aggravate the covid-19 pandemic? early findings and lessons for planners. J. Am. Plan. Assoc. 86, 495–509 (2020).

10. Carozzi, F. Urban density and covid-19. (2020).

11. Keogh-Brown, M. R., Jensen, H. T., Edmunds, W. J. & Smith, R. D. The impact of covid-19, associated behaviours and policies on the uk economy: A computable general equilibrium model. SSM-population health 12, 100651 (2020).

12. Thomsen, P. Transcript of April 2020 European Department Press Briefing. (2020). [Online; accessed 21-January-2021].

13. Wagenaar, A. C., Maybee, R. G. & Sullivan, K. P. Mandatory seat belt laws in eight states: A time-series evaluation. J. Saf. Res. 19, 51–70 (1988).

14. Dennis, J., Ramsay, T., Turgeon, A. F. & Zarychanski, R. Helmet legislation and admissions to hospital for cycling related head injuries in canadian provinces and territories: interrupted time series analysis. Bmj 346 (2013).

15. Hawton, K. et al. Long term effect of reduced pack sizes of paracetamol on poisoning deaths and liver transplant activity in england and wales: interrupted time series analyses. Bmj 346 (2013).

16. Grundy, C. et al. Effect of 20 mph traffic speed zones on road injuries in london, 1986-2006: controlled interrupted time series analysis. Bmj 339 (2009).

17. Lopez Bernal, J. A., Gasparrini, A., Artundo, C. M. & McKee, M. The effect of the late 2000s financial crisis on suicides in spain: an interrupted time-series analysis. The Eur. J. Public Heal. 23, 732–736 (2013).

18. Tran, T. H., Sasikumar, S. N., Hennessy, A., O’Loughlin, A. & Morgan, L. Associations between restrictions on public mobility and slowing of new covid-19 case rates in three countries. The Med. J. Aust. 213, 471 (2020).

19. Armstrong, D. A., Lebo, M. J. & Lucas, J. Do covid-19 policies affect mobility behaviour? evidence from 75 canadian and american cities. Can. Public Policy 46, S127–S144 (2020).

20. Abouk, R. & Heydari, B. The immediate effect of covid-19 policies on social-distancing behavior in the united states. Public Heal. Reports 0033354920976575 (2020).

21. Nguyen, T. D. et al. Impacts of state reopening policy on human mobility. Tech. Rep., National Bureau of Economic Research (2020).

22. Badr, H. S. et al. Association between mobility patterns and covid-19 transmission in the usa: a mathematical modelling study. The Lancet Infect. Dis. 20, 1247–1254 (2020).

23. Badr, H. S. & Gardner, L. M. Limitations of using mobile phone data to model covid-19 transmission in the usa. The Lancet Infect. Dis. (2020).

24. Gatalo, O., Tseng, K., Hamilton, A., Lin, G. & Klein, E. Associations between phone mobility data and covid-19 cases. The Lancet Infect. Dis. (2020).

25. Xiong, C., Hu, S., Yang, M., Luo, W. & Zhang, L. Mobile device data reveal the dynamics in a positive relationship between human mobility and covid-19 infections. Proc. Natl. Acad. Sci. 117, 27087–27089 (2020).

26. Jia, J. S. et al. Population flow drives spatio-temporal distribution of covid-19 in china. Nature 582, 389–394 (2020).

27. Chang, S. et al. Mobility network models of covid-19 explain inequities and inform reopening. Nature 1–6 (2020).

28. Zhou, Y. et al. Effects of human mobility restrictions on the spread of covid-19 in shenzhen, china: a modelling study using mobile phone data. The Lancet Digit. Heal. 2, e417–e424 (2020).

29. Scott, S. L. & Varian, H. R. Predicting the present with bayesian structural time series. Int. J. Math. Model. Numer. Optimisation 5, 4–23 (2014).

30. Brodersen, K. H. et al. Inferring causal impact using bayesian structural time-series models. The Annals Appl. Stat. 9, 247–274 (2015).

